# Leukocyte DNA methylation-based signatures for atherosclerotic cardiovascular disease risk prediction in the Million Veteran Program

**DOI:** 10.64898/2026.08.14.26359154

**Authors:** Alexa Barad, Dennis Khodasevich, Pik Fang Kho, Rodrigo Guarischi-Sousa, Jiayan Zhou, Austin T. Hilliard, Tetsushi Nakao, Pradeep Natarajan, VA Million Veteran Program, Julie A. Lynch, Kyong-Mi Chang, Philip S. Tsao, Andres Cardenas, Shoa L. Clarke, Karen N. Conneely, Yan V. Sun, Themistocles L. Assimes

## Abstract

**Background and Aims:** The contribution of DNA methylation signatures to atherosclerotic cardiovascular disease (ASCVD) risk prediction remains unclear. We developed methylation risk scores (MRS) for incident ASCVD and assessed whether they improved risk prediction beyond established risk factors.

**Methods:** We studied 44,674 Million Veteran Program participants with leukocyte DNA methylation data, divided into two independent subcohorts: a prevalent ASCVD cohort (n=27,560) used for epigenome-wide association analyses (EWAS) to inform cytosine-phosphate-guanine dinucleotide selection, and a cohort free of ASCVD at blood draw (n=17,114), split into training and testing sets for MRS development and evaluation. MRS for incident ASCVD were developed using elastic net regression. Incremental prediction beyond clinical risk factors was assessed by improvement in discrimination (ΔCPE), reclassification (NRI), and calibration.

**Results:** Three MRS were developed: MRS-1A, informed by prevalent ASCVD EWAS and probe reliability; MRS-1B, informed by EWAS alone; and MRS-2, using an agnostic probe reliability-based approach. Among 17,114 participants (mean [SD] age, 58.9 [14.1] years; 89.6% men; 54.2% European), 2,789 developed ASCVD over a median follow-up of 7.4 years. Each MRS was associated with incident ASCVD (HR per 1-SD: 1.97 [95% CI, 1.72-2.26] for MRS-1A, 2.08 [1.83-2.37] for MRS-1B, and 2.07 [1.78-2.39] for MRS-2) and modestly improved discrimination beyond clinical risk factors (ΔCPE: 0.014 [0.006, 0.021], 0.016 [0.007, 0.023], and 0.013 [0.006, 0.021], respectively). MRS improved risk stratification, driven by the downward reclassification of non-events (non-event NRI: 3.6% [2.6-4.7], 5.4% [4.3-6.5], and 3.6% [2.6-4.6], respectively), while maintaining calibration.

**Conclusions:** DNA methylation-based signatures were associated with incident ASCVD and modestly improved risk prediction beyond that of traditional risk factors.

**Key points:** *Key Question:* Do DNA methylation-based risk scores improve the prediction of incident atherosclerotic cardiovascular disease (ASCVD) beyond the established clinical risk factors?

*Key Finding:* DNA methylation-based risk scores were strongly associated with incident ASCVD and provided modest but statistically significant improvement in risk prediction beyond traditional clinical risk factors in a primary prevention cohort.

*Take-home Message:* These findings suggest that leukocyte DNA methylation-based signatures may provide incremental value for ASCVD risk assessment and support the further evaluation of epigenetic biomarkers in multi-omics risk-prediction frameworks.

## Introduction

Cardiovascular disease is the leading cause of morbidity and mortality worldwide. Atherosclerotic cardiovascular disease (ASCVD), including coronary artery disease (CAD), peripheral artery disease (PAD), and ischemic stroke, accounts for most of the global cardiovascular disease burden and was responsible for more than 12 million deaths in 2023.^1^ Because ASCVD develops gradually over decades, clinical disease often manifests only after substantial subclinical pathology has occurred.^2^ Therefore, early identification of individuals at elevated risk is essential for effective disease prevention.

Clinical risk prediction models play a central role in the prevention of cardiovascular diseases by guiding the initiation and intensity of preventive therapies. Contemporary risk assessment integrates established risk factors, such as age, sex, blood pressure, cholesterol levels, smoking status, and diabetes.^3–5^ Although these risk factors account for a substantial proportion of ASCVD risk, there is considerable interest in identifying complementary biomarkers that further enhance risk prediction. However, demonstrating meaningful incremental improvements has proven challenging because the traditional risk factors already provide strong predictive information.^6,7^ Numerous studies have evaluated molecular biomarkers, including genomic,^8–10^ proteomic,^11–13^ and metabolomic predictors,^14–16^ yet improvements over traditional risk factors have generally been modest. Because ASCVD arises from the combined effects of genetic susceptibility, environmental exposures, and behavioral factors across the life course, integrating complementary molecular layers may improve cardiovascular risk prediction.^17^ In addition to genomics, proteomics, and metabolomics, epigenetic biomarkers represent another molecular layer that remains understudied in ASCVD risk prediction.

DNA methylation is a key epigenetic modification that reflects both environmental exposures and downstream biological responses.^18^ Methylation patterns can capture cumulative exposures related to smoking, diet, metabolic dysfunction, and inflammation, which are central to the development of atherosclerosis.^19,20^ Epigenome-wide association studies (EWAS) have identified multiple loci associated with cardiovascular risk factors and incident cardiovascular disease.^20–24^ However, few studies have evaluated whether DNA methylation signatures improve ASCVD risk prediction in primary prevention settings, and most have been limited by substantially smaller sample sizes than studies of other omics layers.^20,25,26^ Therefore, the potential contribution of epigenetic biomarkers to ASCVD risk prediction remains unclear.

In this study, we leveraged large-scale multi-ancestry genome-wide DNA methylation data from the Million Veteran Program (MVP) to develop and evaluate methylation-based risk scores (MRS) for incident ASCVD and to assess whether these epigenetic biomarkers provide information beyond established risk factors. These analyses may help inform future efforts to integrate epigenetic biomarkers into multi-omics-based ASCVD risk prediction models.

## Methods

### Study Population

The MVP, initiated in 2011, is a nationwide, multi-ancestry observational cohort study designed to evaluate how genetic, lifestyle, and environmental factors influence health and disease risk among U.S. Veterans. Participants provided peripheral blood samples for DNA extraction and multi-omics profiling, including genotyping, whole-genome sequencing, and other molecular assays, and consented to access their Veterans Health Administration (VHA) electronic health record (EHR) both before and after enrollment. Participants were also asked to complete optional baseline and lifestyle questionnaires to complement the EHR data. The MVP received ethical and study protocol approval from the VA Central Institutional Review Board, and all the participants provided informed consent. Detailed descriptions of the study design and recruitment procedures have been published.^27^

The DNA methylation subsample consisted of 45,446 individuals derived from the genotyped MVP cohort (R4) through a combination of random sampling and targeted enrichment for participants with post-traumatic stress disorder (PTSD), chronic kidney disease (CKD), PAD, extreme hypertension, or those aged > 80 years at enrollment. For risk prediction analyses, we created a pseudorandom incident subset of participants free of ASCVD at baseline (incident cohort, n = 17,114 participants). Sampling fractions derived from the full MVP genotyped cohort, based on age categories (≤ 40, 41-60, 61-80, > 80 years), PTSD, CKD, and diabetes, were applied to randomly downsample overrepresented strata, thereby improving the approximation to the broader MVP population. Separately, we generated an independent subset for the prevalent ASCVD EWAS, including prevalent ASCVD cases and randomly selected non-cases at blood draw in a 1:1 ratio (prevalent cohort, n = 27,560 participants). A detailed flowchart illustrating the generation of these incident and prevalent cohorts is shown in **Figure S1**.

### DNA methylation

DNA methylation was measured using an Illumina Infinium MethylationEPIC BeadChip array (EPIC v1). Raw data were processed in R using the *SeSAMe* package (v1.8.2).^28^ Probe-level quality control excluded probes that failed detection using the Out-Of-Band Array Hybridization (pOOBAH) method, defined as a detection *P*-value > 0.05 in ≥ 10% of samples.^29^ At the sample level, individuals with a probe pass rate < 96% were excluded. We further removed technically unreliable probes (including those with poor mapping quality, poor titration correlation, or evidence of color channel switching), SNP-associated probes identified in the Illumina manifest, and probes located on sex chromosomes. A total of 721,647 probes remained for analysis. Blood cell-type proportions (CD4T, CD8T, B cells, Monocytes, Eosinophils, Neutrophils, and NK cells) were calculated with the Houseman method.^30^ Further details on quality control and preprocessing procedures have been described previously.^31^

### Genotyping

Genotyping data were obtained using a custom ThermoFisher Axiom MVP 1.0 array, as previously described,^32^ and further expanded to the current fourth release (R4). Genetic ancestry was inferred using the 1000 Genomes Project reference panel, with each participant assigned to one of five superpopulations: African (AFR), Admixed American (AMR), East Asian (EAS), European (EUR), or South Asian (SAS). Participants who could not be confidently assigned to any of these groups were classified as “Undetermined.” Genetic principal components were calculated across the entire cohort and included as covariates in the analyses.

### Atherosclerotic cardiovascular disease definition

ASCVD was defined as a composite of CAD, PAD, acute ischemic stroke (AIS), and death related to ASCVD. Prevalent cases were those with a first documented diagnosis on or before the blood draw date, and incident cases were those diagnosed afterward. CAD cases were identified using diagnostic and procedural codes from the VHA or Medicare hospital records. Participants were classified as cases if they had at least one procedural code indicating acute myocardial infarction or coronary revascularization. In the absence of procedural codes, individuals with at least two diagnostic codes for CAD recorded within one year, provided that they were not clustered within a two-month period, were also classified as cases. PAD cases were defined by any of the following: two or more PAD diagnoses on separate dates, a PAD diagnosis as the primary inpatient diagnosis, a PAD diagnosis with a lower extremity revascularization or amputation procedure, a PAD diagnosis accompanied by two or more ankle-brachial index tests within 14 months, or a PAD diagnosis accompanied by two or more vascular surgery clinic visits within 14 months.^33^ AIS cases were identified using an algorithm based on diagnostic codes as described previously.^34^ ASCVD-related deaths were defined as deaths for which any of the following ICD-10 codes were listed as the primary cause in the National Death Index: I10, I11, I13, I16, I20–I25, I46, I63, I67, I70, I74, I75, and G45.^8^ Participants who did not meet the criteria for CAD, PAD, AIS, or ASCVD-related death were considered non-cases.

For survival analyses, the time-to-ASCVD was defined as the interval from the blood draw date to the first ASCVD diagnosis. In participants with multiple ASCVD events, the earliest event date was selected. For analyses of ASCVD subtypes, the time-to-event was calculated separately for each outcome, allowing participants with multiple ASCVD events to contribute to multiple subtype analyses, thereby maximizing the case sample sizes. For non-cases, the follow-up time was calculated from the blood draw to the last VHA visit, the end of follow-up (January 8, 2024), or non-ASCVD death, whichever occurred first. Non-cases with a recorded last VHA visit date preceding their blood draw were excluded from the survival analyses (n = 203; n = 70 in the test set only).

### Demographic and clinical variables

Participant age and sex were obtained from the MVP baseline questionnaires, with age defined at the time of blood collection. The body mass index (kg/m²) was calculated from height and weight recorded in the VHA EHR or, when unavailable, from the baseline questionnaire data. Smoking status (never, former, or current) was determined using a validated algorithm developed from VHA EHR data, as previously described.^35^ Medication use during the three years prior to enrollment was identified using VA National Formulary drug classes and categorized as diabetes medications (insulin, oral hypoglycemic agents, and glucagon-like peptide-1 receptor agonists), lipid-lowering agents (antilipemic agents), and antihypertensive medications (beta-blockers, calcium channel blockers, diuretics, and renin-angiotensin system inhibitors). Laboratory measurements and vitals, including high-density lipoprotein cholesterol (HDL-C), low-density lipoprotein cholesterol (LDL-C), and systolic blood pressure, were extracted from EHR records using values obtained within two years prior to enrollment. Prevalent diabetes at the time of blood draw was defined as ≥1 inpatient or primary care encounter with ICD-9 code 250. xx or ICD-10 codes E08.x, E09.x, E10.x, E11.x, or E13.x, or ≥2 occurrences of these codes in any clinical setting, in addition to an outpatient prescription for a diabetes medication based on the VA National Formulary drug classes. Histories of hypertension, PTSD, and CKD at the time of blood draw were defined using phecodes derived from EHR data (hypertension: phecode 401; PTSD: phecode 300.9; CKD: phecodes 585.3 and 585.4).

### Statistical Analyses

**Figure 1** summarizes the study design and analytical workflow. To address the computational challenges of modeling genome-wide DNA methylation in penalized regression, cytosine-phosphate-guanine dinucleotide (CpG) sites were pre-filtered using an EWAS strategy, followed by complementary filtering approaches, as described below.

**Figure 1.**
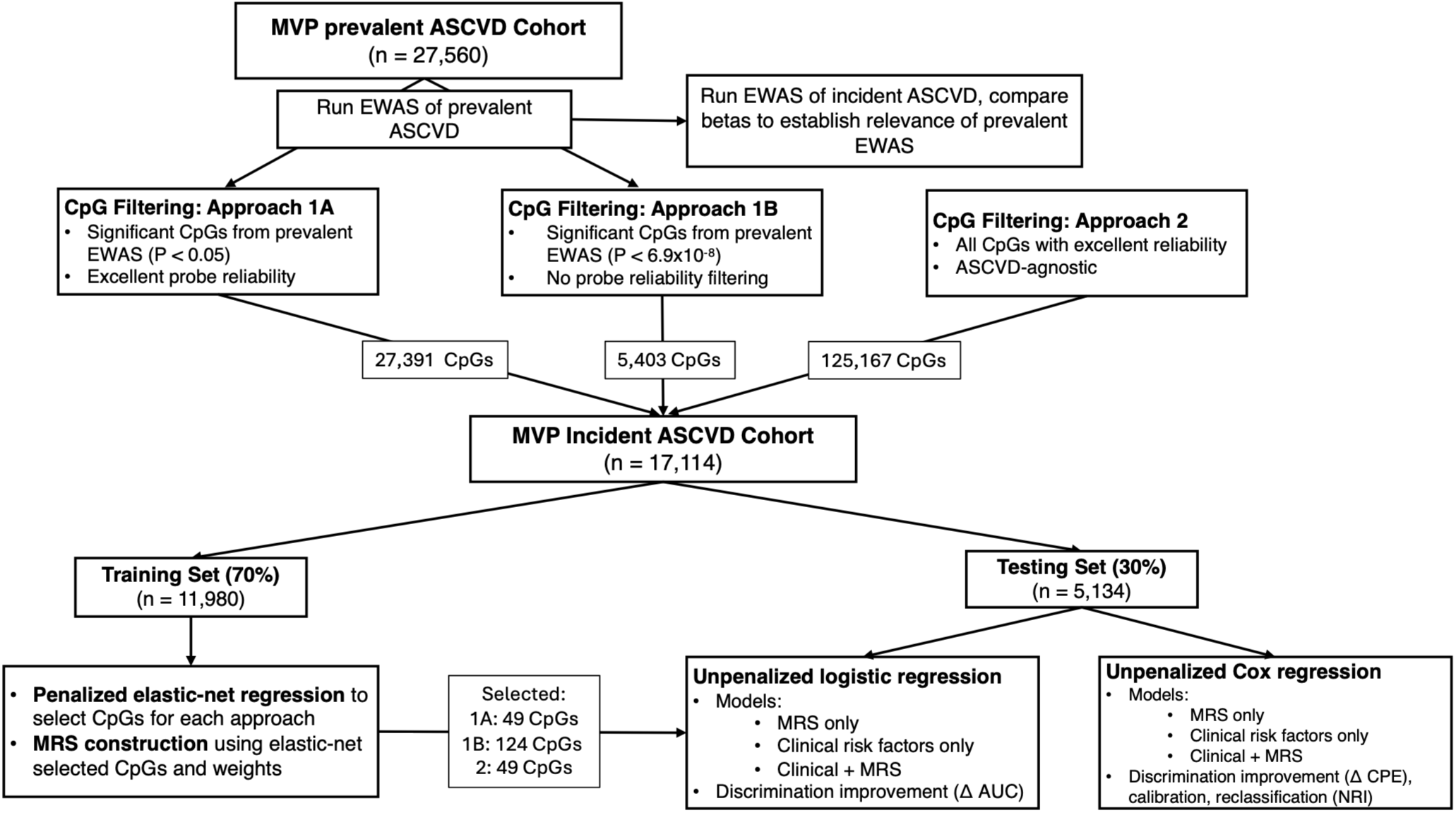
Study design and analytical pipeline. The prevalent and incident cohorts were completely independent. ASCVD, atherosclerotic cardiovascular disease; EWAS, Epigenome-wide association study; MRS, methylation risk score.

### EWAS for CpG pre-filtering

In the independent prevalent cohort (n = 27,560), we conducted an EWAS of prevalent ASCVD using DNA methylation beta values and the *Meffil* R package (v1.3.4).^36^ Models were adjusted for age, age², sex, estimated blood cell proportions, the first 20 genetic principal components, the first 20 technical principal components, DNA methylation scanner, and sample storage duration. To control for bias and genomic inflation, we applied the *bacon* R package (v1.31.1),^37^ which implements a Bayesian adjustment method. Statistical significance was defined using a Bonferroni-corrected threshold of 6.9×10^−8^ (0.05/721,647 probes tested) applied to the bacon-adjusted *P*-values. To assess the prospective relevance of CpGs identified in the prevalent ASCVD EWAS, we conducted an additional EWAS of incident ASCVD in an independent cohort (n = 17,114) using identical regression models. Relevance was evaluated based on the overlap of significant CpGs and concordance of effect estimates between the prevalent and incident EWAS. Results from the incident EWAS were used solely for validation and were not incorporated into downstream feature selection to preserve the independence between cohorts.

Using Bonferroni-significant CpGs from the prevalent ASCVD EWAS as input, we performed Kyoto Encyclopedia of Genes and Genomes (KEGG) pathway enrichment analysis using the *gometh* function in the missMethyl R package (v1.36.0)^38^ and Reactome pathway enrichment analysis using ReactomePA R package (v1.46.0).^39^

### CpG filtering strategies

Three complementary filtering strategies were applied to define the CpG sets for modeling. Approaches 1A and 1B leveraged the results of the prevalent ASCVD EWAS. Approach 1A included CpGs meeting a nominal significance level (*P* < 0.05) and restricted to probes with excellent measurement reliability (intraclass correlation coefficients > 0.75 based on duplicate samples).^40^ Approach 1B included all CpGs surpassing the Bonferroni-corrected threshold (*P* < 6.9×10^−8^, 0.05/721,647 probes tested) without further filtering. Approach 2 included all probes with excellent reliability,^40^ independent of prior associations.

### Methylation risk scores construction and internal validation

Incident ASCVD analyses were conducted in an independent incident cohort free of ASCVD at blood draw (n = 17,114), which was randomly split 70/30 into training (n = 11,980) and testing (n = 5,134) sets. Elastic net regression with α = 0.5 and 10-fold cross-validation to select the λ whose cross-validation error was within one standard error of the minimum (λ_1SE) was applied separately to each CpG set (1A, 1B, and 2). The retained CpGs were used to construct the MRS as weighted sums of methylation beta values, using elastic net coefficients as weights, resulting in three scores corresponding to the filtering approaches (MRS-1A, MRS-1B, and MRS-2).

The initial model performance was evaluated in the testing set using logistic regression with 1) MRS only, 2) clinical risk factors only, and 3) clinical risk factors plus MRS. Clinical risk factors were selected to reflect contemporary clinical risk assessment,^5^ including body mass index, smoking, HDL-C, LDL-C, systolic blood pressure, lipid-lowering medication use, antihypertensive medication use, and diabetes history. Incremental prediction was assessed by comparing the AUC between the clinical-only and clinical-plus-MRS models, with statistical significance evaluated using the DeLong test.^41^ Improvement in model fit by adding MRS beyond clinical risk factors was evaluated using likelihood ratio tests (LRT). All models were adjusted for age, sex, genetic principal components 1-10, and estimated blood cell proportions.

### Prospective analyses of incident ASCVD

The prospective associations of MRS with incident ASCVD were assessed using Cox regression in the test set only, with results presented as HR [95%CI] and discrimination quantified by concordance probability estimate (CPE). The incremental predictive value was calculated as ΔCPE between the clinical-only and clinical-plus-MRS models, with 95% CIs estimated via bootstrapping. The proportional hazards assumption was evaluated using Schoenfeld residuals and the *cox.zph* R function, and no violations were detected. The MVP-derived ASCVD MRS (1A, 1B, 2) were compared with previously published CAD-specific MRS (MRS-Westerman_ComBat and MRS-Westerman_combined).^25^ We additionally evaluated all MRS against the DNA methylation-based biomarker of mortality, GrimAge2.^42^ GrimAge2 was selected as the benchmark epigenetic clock as it has demonstrated the strongest associations with cardiovascular disease outcomes.^43–45^ MRS-Westerman_ComBat, MRS-Westerman_combined, and GrimAge2 were calculated using published weights.^25,42,43^ Improvement in model fit by adding the MRS beyond clinical risk factors, GrimAge2, or clinical risk factors plus GrimAge2 was evaluated with LRT. All MRS and GrimAge2 measures were standardized (mean = 0, SD = 1), with HR [95%CI] representing the risk per 1-SD increase.

For each MRS developed, the categorical net reclassification improvement (NRI) was estimated using the *nricens* R package (v1.6) at 5-and 10-year horizons. The 10-year horizon was selected for clinical relevance; 5-year estimates were also reported, given that only ∼20% of participants had follow-up ≥ 10 years. Risk categories were defined using clinically aligned thresholds of 5% and 10% for the 10-year risk^46^ and scaled to 2.5% and 5% for the 5-year risk, respectively. Calibration-in-the-large was assessed as the difference between the observed event rate and the mean of predicted probabilities based on Cox modeling,^47^ and calibration plots were generated using the *Score* and *plotCalibration* functions from the *riskRegression* R package (v2023.12.21) at both time horizons.

### Sensitivity analyses

To assess potential misclassification arising from the inclusion of ASCVD deaths identified via the National Death Index, we conducted sensitivity analyses using an alternative ASCVD definition that excluded these deaths. Methylation risk scores were re-derived in the training set using this modified outcome definition and subsequently evaluated in the independent test set using the same modeling framework as in the primary analyses.

## Results

### Study cohort

The total analytic sample included 44,674 participants representing the MVP subset with methylation data (**Table 1**). The mean age of the total sample was 64.5 years (SD, 14.3); 93% were men, 60.4% were of European ancestry, 26.5% were current smokers, and 30.7% had a history of diabetes. The prevalence of oversampled conditions was 20.3% for PAD, 9.2% for CKD, and 14.7% for PTSD.

**Table 1.**
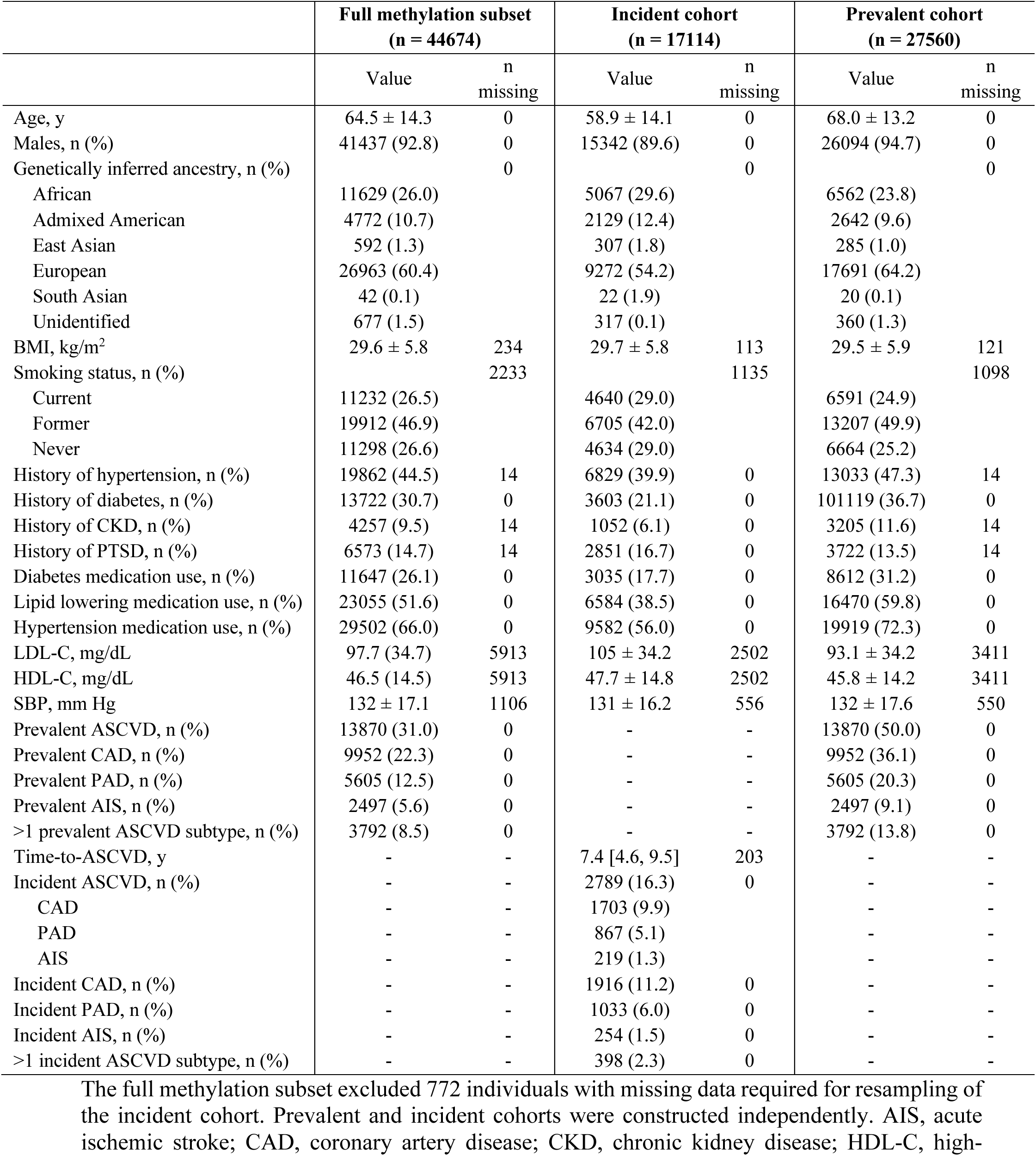
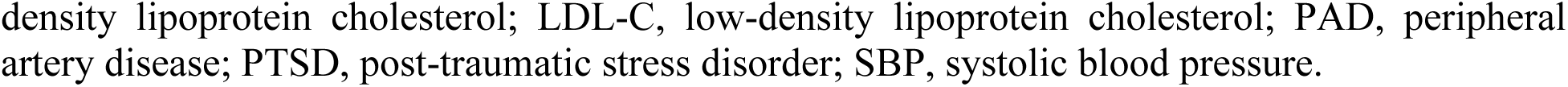
Descriptive characteristics of the full MVP DNA methylation subset and prevalent and incident cohorts.

For risk prediction analyses, a pseudorandom subset of 17,114 participants free of ASCVD at blood draw comprised the incident cohort after selective downsampling of specific groups (**Table 1**). Following downsampling, the incident cohort had a mean age of 58.9 years (SD, 14.1), and a prevalence of 21.1% for diabetes, 6.2% for CKD, and 16.7% for PTSD, closely matching those of the full MVP genotyped cohort (**Table S1**). During a median follow-up of 7.4 years, 2,789 (16.3%) incident ASCVD events occurred, including 1,916 CAD, 1,033 PAD, and 254 AIS, with 398 participants experiencing more than one ASCVD subtype during follow-up. The high incidence of ASCVD likely reflects the non-normal age distribution of Veterans at enrollment, with over 50% of the cohort aged 60 to 80 years, combined with the high burden of risk factors among Veterans.^27^ The characteristics of the incident cohort stratified by the training and testing sets are shown in **Table S2.**

The prevalent cohort comprised 27,560 participants, including 13,870 with ASCVD at the time of blood collection. As expected, given the sampling approach, individuals in the prevalent cohort were generally older (mean, 68.0; SD, 13.2 years) and had a higher prevalence of cardiometabolic comorbidities than those in the incident cohort (**Table 1**). The characteristics of the prevalent ASCVD cohort stratified by case status are shown in **Table S3**. Among prevalent cases, the mean age at first ASCVD diagnosis was 64.7 years (SD, 10.5), with a median of 6.6 years between the first diagnosis and the blood sample collection.

### EWAS of ASCVD for CpG filtering

Summary statistics and annotations for all Bonferroni-significant CpG associations from the prevalent and incident EWAS are provided in **Supplementary Material 2.** Miami and quantile-quantile plots for both EWAS are shown in **Figures S2-S3**. A total of 5,403 and 101 CpG sites in the prevalent and incident ASCVD EWAS, respectively, surpassed the Bonferroni-corrected significance threshold. Effect estimates for CpG sites that were significant in at least one EWAS were strongly correlated (r = 0.93) (**Figure S4**). There was also substantial overlap in the differentially methylated sites identified across the EWAS, with 93% of CpGs reaching Bonferroni significance in the incident EWAS also identified in the prevalent EWAS (**Figure S4**). Together, the strong correlation and high degree of overlap support the prospective relevance of CpGs identified in the prevalent ASCVD EWAS.

Kyoto Encyclopedia of Genes and Genomes pathway enrichment analysis of genes annotated to Bonferroni-significant CpG sites from the prevalent ASCVD EWAS identified broad inflammatory pathways, including chemokine signaling, tumor necrosis factor signaling, and apoptosis (**Table S4**). Pathways such as non-small cell lung carcinoma also appeared enriched, possibly reflecting the strong influence of smoking-associated CpG sites (**Table S4)**. Complementary Reactome analysis highlighted biological processes relevant to atherosclerosis, including platelet activation, vascular-endothelial growth factor signaling, sterol regulatory element-binding protein-mediated regulation of cholesterol homeostasis, and peroxisome proliferator-activated receptor α-mediated lipid metabolism (**Figure S5**).

### Development and internal validation of the ASCVD MRS

The CpG filtering strategies yielded 27,391, 5,403, and 125,167 CpGs for approaches 1A, 1B, and 2, respectively, from which the elastic net models selected 49, 124, and 49 CpGs. The selected CpGs and corresponding elastic net coefficients used to compute each MRS are provided in **Supplementary Material 3**. The three MRS were strongly correlated with each other, with all pairwise correlations between 0.94 and 0.99. In the independent test set, the prediction of incident ASCVD using each MRS produced AUC values ranging from 0.705 to 0.717 across approaches (**Figure 2**). An unpenalized logistic regression model including clinical risk factors alone achieved an AUC of 0.709, and adding MRS significantly improved discrimination for all approaches, with AUC increases of 0.011 to 0.013 (DeLong test, *P* < 0.05) (**Figure 2**). Consistent with these findings, the LRT indicated a significant improvement in the model fit when each MRS was added to the clinical risk factor model (all LRT P<2×10^−16^).

**Figure 2.**
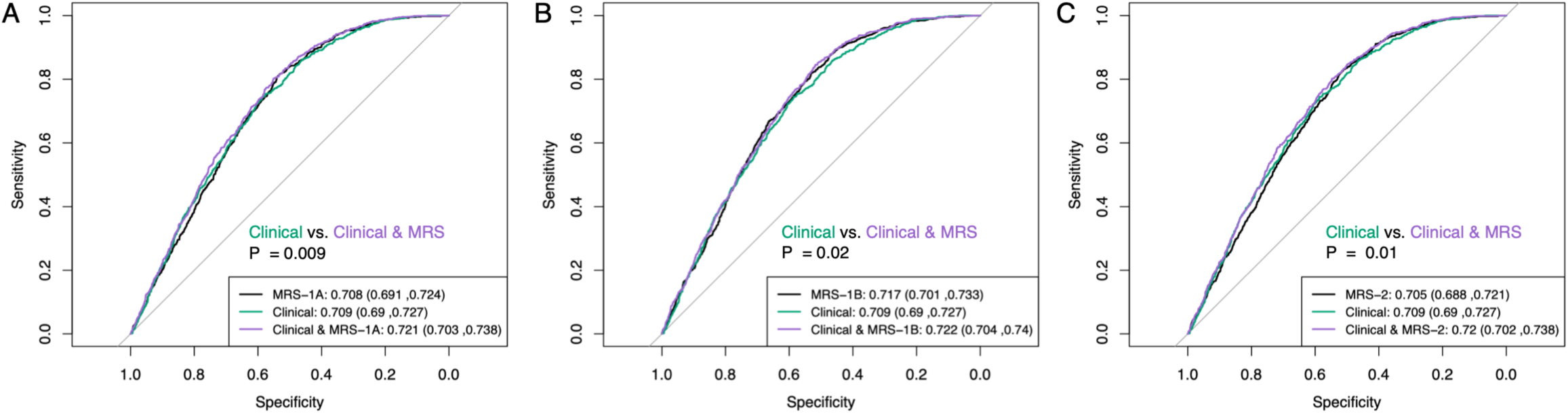
Discrimination of elastic net-derived methylation risk scores (MRS) for ASCVD. Receiver operating characteristic curves from logistic regression models comparing MRS-only models, clinical risk factors alone, and combined clinical + MRS models across the three elastic net approaches (Panels A-C). AUC values (95% CI) are shown for each model. P-values reflect comparisons of the clinical and clinical + MRS models using DeLong’s test. Analyses were conducted using the held-out test set (n = 5,134 for MRS-only models; n = 4,172 for clinical and clinical + MRS models). The clinical models included body mass index, smoking status, HDL-C, LDL-C, systolic blood pressure, lipid-lowering medication use, antihypertensive medication use, and diabetes history. All models were adjusted for age, sex, genetic principal components 1–10, and estimated blood cell proportions.

To assess the robustness of the MRS, we evaluated the dependence on training-test splits using nested 10-fold cross-validation, which demonstrated stable discrimination across folds (**Figure S6**). We further examined the robustness to alternative elastic net specifications by varying the penalization parameters, including α (0.1-1.0) and λ (λ_1SE and λ_min) (**Table S5**). Across these specifications, the model with α = 0.5 and λ_1SE consistently achieved the most favorable balance between discrimination and parsimony. Although less-penalized models (e.g., λ_min or smaller α) selected substantially more CpGs (>300) and yielded modestly higher AUCs in MRS-only models, these gains were not retained after integration with clinical risk factors, suggesting potential overfitting and limited incremental value of larger CpG sets in this study.

Sensitivity analyses using an ASCVD definition excluding deaths yielded consistent results. MRS re-derived under this definition demonstrated similar improvements in the prediction of incident ASCVD (excluding deaths), with AUC increases over clinical risk factors ranging from 0.011 to 0.015 (DeLong test, *P* < 0.05) (**Figure S7**).

### Prospective association and comparative performance of the ASCVD MRS

Prospective associations between each MRS and incident ASCVD were evaluated in the independent test set with complete time-to-event and clinical covariate data (n = 4,172; 725 cases). Analyses were conducted for each MVP-derived MRS as well as for two previously published CAD-specific MRS (MRS-Westerman_ComBat and MRS-Westerman_combined).^25^ Correlations between the MVP-derived and previously published CAD-specific MRS ranged from 0.84 to 0.87.

Kaplan-Meier plots of the time to incident ASCVD by tertiles of each MRS are shown in **Figure 3**, with corresponding plots for ASCVD subtypes shown in **Figure S8**. Across the three MVP MRS, higher scores were consistently associated with an increased risk of incident ASCVD, with HRs per 1-SD of 1.97 (95% CI, 1.72-2.26) for MRS-1A, 2.08 (95% CI, 1.83-2.37) for MRS-1B, and 2.07 (95% CI, 1.78-2.39) for MRS-2 (**Figure 4A**). Similar, though smaller, associations were observed for the previously published CAD-specific scores, with HRs of 1.60 (95% CI, 1.43-1.78) for MRS-Westerman_combined and 1.52 (95% CI, 1.37-1.69) for MRS-Westerman_ComBat (**Figure 4A**), consistent with effect sizes reported in the original study (HRs ranging from 1.28 to 1.58).^25^ All five MRS remained significantly associated with incident ASCVD after adjusting for clinical risk factors (**Figure 4A**). In disease-stratified analyses (**Figures 4B-D**), the associations for CAD were similar between the MVP MRS and the previously published CAD-specific MRS (**Figure 4B**), whereas the MVP MRS showed larger effect estimates for PAD than the CAD-specific MRS (**Figure 4C**). The associations with AIS were generally consistent but were estimated with wider confidence intervals, as expected, given the substantially smaller case count (**Figure 4D**). The results were consistent when the ASCVD definition was used without ASCVD deaths (**Figure S9**).

**Figure 3.**
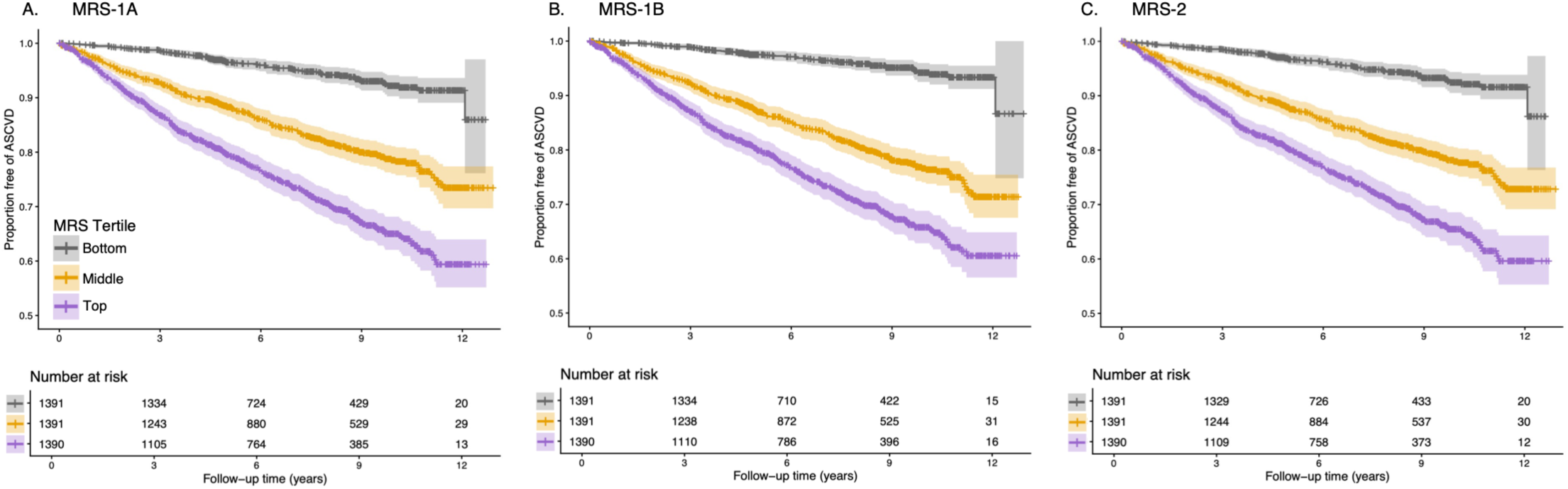
Kaplan-Meier curves for incident ASCVD stratified by methylation risk score (MRS) tertiles. Kaplan-Meier estimates of ASCVD-free survival across tertiles of the methylation risk score derived from the three elastic net approaches: MRS-1A (Panel A), MRS-1B (Panel B), and MRS-2 (Panel C). Participants were categorized into tertiles corresponding to the bottom, middle, and top thirds of the MRS distributions. The shaded areas represent the 95% confidence intervals.

**Figure 4.**
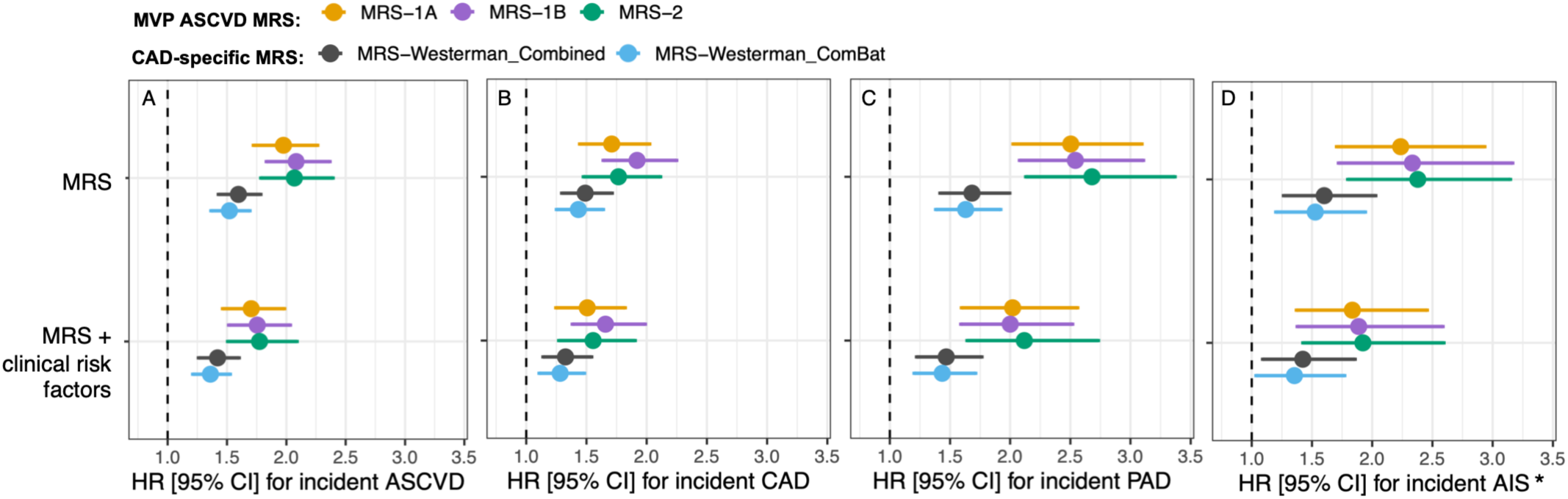
Associations between methylation risk scores (MRS) and incident ASCVD and ASCVD subtypes. Hazard ratios (HRs) and 95% confidence intervals from Cox proportional hazards models for incident ASCVD (A), coronary artery disease (CAD) (B), peripheral artery disease (PAD) (C), and acute ischemic stroke (AIS) (D). The number of cases was 725 for ASCVD, 489 for CAD, 285 for PAD, and 67 for AIS in this study. HR [95%CI] are shown per 1-SD increase in each MRS. The top row shows models including the MRS alone, and the bottom row shows models additionally adjusted for clinical risk factors, including body mass index, smoking status, HDL-C, LDL-C, systolic blood pressure, lipid-lowering medication use, antihypertensive medication use, and diabetes history. All models were adjusted for age, sex, genetic principal components 1-10, and estimated blood cell proportions. *\**Due to the smaller number of AIS cases, models for AIS were adjusted for a reduced set of base covariates (age, genetic principal components 1-5, and estimated blood cell proportions).

Next, we evaluated whether the inclusion of the MRS improved prediction beyond clinical risk factors (**Table 2**). The clinical risk factor model alone yielded a CPE of 0.699 (95% CI, 0.681-0.716). Among the MRS-only models, MRS-1B demonstrated discrimination comparable to that of the clinical risk factor model (CPE, 0.696 vs. 0.699), whereas MRS-1A (CPE, 0.688) and MRS-2 (CPE, 0.687) yielded slightly lower discrimination. Addition of the MVP MRS to the clinical risk factor model modestly improved discrimination, increasing CPE from 0.699 to 0.712-0.715 (ΔCPE, 0.013-0.016). Similar but slightly smaller improvements were observed for the previously published CAD-specific scores, with CPE values of 0.706-0.708 (ΔCPE, 0.007-0.009). The inclusion of each MRS significantly improved the model fit beyond clinical risk factors alone, according to the LRT (all LRT P< 1×10^−6^). In disease-stratified prediction analyses, the inclusion of MRS modestly improved discrimination for CAD and PAD, with smaller, non-significant improvements for AIS (**Table 3**). For PAD, among the MRS-only models, MRS-1B demonstrated better discrimination than the clinical risk factor model (CPE, 0.743 vs. 0.738, respectively), and the addition of MRS to the clinical risk factor model produced the greatest improvement in discrimination for this outcome (ΔCPE, 0.020–0.022). These findings may reflect the greater severity of the disease or the ability of DNA methylation signatures to capture the cumulative effects of exposures relevant to PAD risk. The findings were similar when ASCVD was defined by excluding ASCVD-related deaths (**Table S6**).

**Table 2.** Incremental predictive value of methylation risk scores (MRS) for incident ASCVD beyond clinical risk factors.

| **MRS** | **Model** | **CPE [95% CI]** | **ΔCPE [95% CI]** | **LRT P-value** |
| --- | --- | --- | --- | --- |
| MVP MRS-1A | **MRS** | 0.688 [0.671, 0.706] | - | - |
|  | **Clinical** | 0.699 [0.681, 0.716] | - | - |
|  | **Clinical + MRS** | 0.713 [0.696, 0.729] | 0.014 [0.006, 0.021] | 1.4e-11 |
| MVP MRS-1B | **MRS** | 0.696 [0.678, 0.713] | - | - |
|  | **Clinical** | 0.699 [0.681, 0.716] | - | - |
|  | **Clinical + MRS** | 0.715 [0.698, 0.731] | 0.016 [0.007, 0.023] | 1.1e-13 |
| MVP MRS-2 | **MRS** | 0.687 [0.670, 0.704] | - | - |
|  | **Clinical** | 0.699 [0.681, 0.716] | - | - |
|  | **Clinical + MRS** | 0.712 [0.695, 0.729] | 0.013 [0.006, 0.021] | 1.2e-11 |
| Westerman<br>MRS-combined<sup>1</sup> | **MRS** | 0.678 [0.660, 0.695] | - | - |
|  | **Clinical** | 0.699 [0.681, 0.716] | - | - |
|  | **Clinical + MRS** | 0.708 [0.691, 0.725] | 0.009 [0.003, 0.015] | 2.4e-8 |
| Westerman<br>MRS-ComBat<sup>1</sup> | **MRS** | 0.673 [0.655, 0.690] | - | - |
|  | **Clinical** | 0.699 [0.681, 0.716] | - | - |
|  | **Clinical + MRS** | 0.706 [0.689, 0.723] | 0.007 [0.002, 0.013] | 3.4e-7 |

CPE and 95% confidence intervals from Cox proportional hazards models for incident ASCVD in the independent test set (n = 4,172; 725 ASCVD cases). The models included MRS alone, clinical risk factors alone, and clinical risk factors plus MRS. ΔCPE represents the difference in CPE between the clinical + MRS and clinical risk factor models. LRT P-values evaluate the improvement in model fit with the addition of the MRS to the clinical model. The clinical models included body mass index, smoking, HDL-C, LDL-C, systolic blood pressure, lipid-lowering medication use, antihypertensive medication use, and diabetes history. All models were adjusted for age, sex, genetic principal components 1-10, and estimated blood cell proportions. ASCVD, atherosclerotic cardiovascular disease; CPE, concordance probability estimate; ΔCPE, change in concordance probability estimate; LRT, likelihood ratio test; MRS, methylation risk score.
<sup>1</sup>Previously published CAD-specific MRS.<sup>25</sup>

**Table 3.**
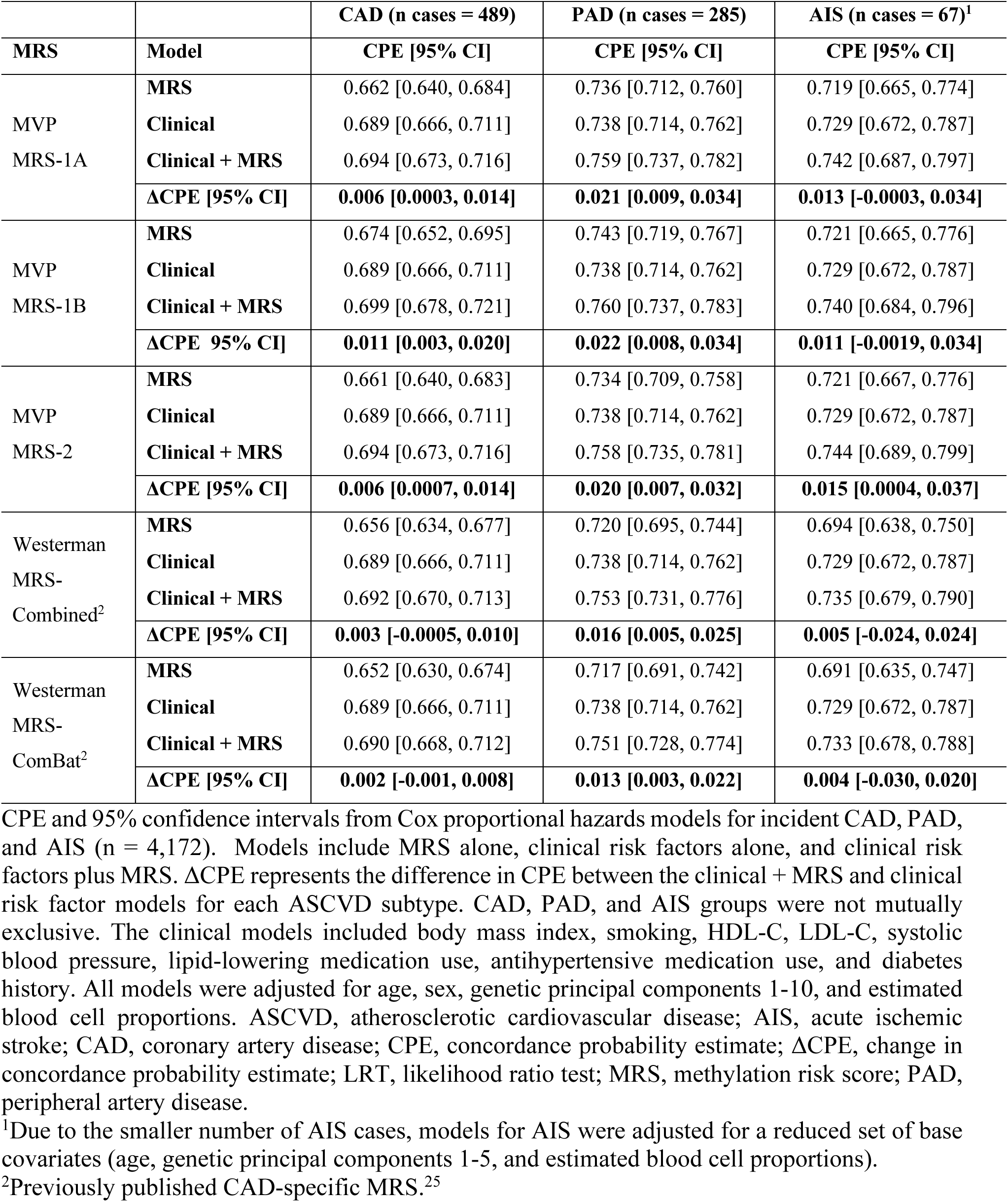
Incremental predictive value of methylation risk scores (MRS) for incident ASCVD subtypes beyond clinical risk factors.

|  |  | **CAD (n cases = 489)** | **PAD (n cases = 285)** | **AIS (n cases = 67)<sup>1</sup>** |
| --- | --- | --- | --- | --- |
| **MRS** | **Model** | **CPE [95% CI]** | **CPE [95% CI]** | **CPE [95% CI]** |
| MVP<br>MRS-1A | **MRS** | 0.662 [0.640, 0.684] | 0.736 [0.712, 0.760] | 0.719 [0.665, 0.774] |
|  | **Clinical** | 0.689 [0.666, 0.711] | 0.738 [0.714, 0.762] | 0.729 [0.672, 0.787] |
|  | **Clinical + MRS** | 0.694 [0.673, 0.716] | 0.759 [0.737, 0.782] | 0.742 [0.687, 0.797] |
|  | **ΔCPE [95% CI]** | **0.006 [0.0003, 0.014]** | **0.021 [0.009, 0.034]** | **0.013 [-0.0003, 0.034]** |
| MVP<br>MRS-1B | **MRS** | 0.674 [0.652, 0.695] | 0.743 [0.719, 0.767] | 0.721 [0.665, 0.776] |
|  | **Clinical** | 0.689 [0.666, 0.711] | 0.738 [0.714, 0.762] | 0.729 [0.672, 0.787] |
|  | **Clinical + MRS** | 0.699 [0.678, 0.721] | 0.760 [0.737, 0.783] | 0.740 [0.684, 0.796] |
|  | **ΔCPE 95% CI]** | **0.011 [0.003, 0.020]** | **0.022 [0.008, 0.034]** | **0.011 [-0.0019, 0.034]** |
| MVP<br>MRS-2 | **MRS** | 0.661 [0.640, 0.683] | 0.734 [0.709, 0.758] | 0.721 [0.667, 0.776] |
|  | **Clinical** | 0.689 [0.666, 0.711] | 0.738 [0.714, 0.762] | 0.729 [0.672, 0.787] |
|  | **Clinical + MRS** | 0.694 [0.673, 0.716] | 0.758 [0.735, 0.781] | 0.744 [0.689, 0.799] |
|  | **ΔCPE [95% CI]** | **0.006 [0.0007, 0.014]** | **0.020 [0.007, 0.032]** | **0.015 [0.0004, 0.037]** |
| Westerman<br>MRS-Combined<sup>2</sup> | **MRS** | 0.656 [0.634, 0.677] | 0.720 [0.695, 0.744] | 0.694 [0.638, 0.750] |
|  | **Clinical** | 0.689 [0.666, 0.711] | 0.738 [0.714, 0.762] | 0.729 [0.672, 0.787] |
|  | **Clinical + MRS** | 0.692 [0.670, 0.713] | 0.753 [0.731, 0.776] | 0.735 [0.679, 0.790] |
|  | **ΔCPE [95% CI]** | **0.003 [-0.0005, 0.010]** | **0.016 [0.005, 0.025]** | **0.005 [-0.024, 0.024]** |
| Westerman<br>MRS-ComBat<sup>2</sup> | **MRS** | 0.652 [0.630, 0.674] | 0.717 [0.691, 0.742] | 0.691 [0.635, 0.747] |
|  | **Clinical** | 0.689 [0.666, 0.711] | 0.738 [0.714, 0.762] | 0.729 [0.672, 0.787] |
|  | **Clinical + MRS** | 0.690 [0.668, 0.712] | 0.751 [0.728, 0.774] | 0.733 [0.678, 0.788] |
|  | **ΔCPE [95% CI]** | **0.002 [-0.001, 0.008]** | **0.013 [0.003, 0.022]** | **0.004 [-0.030, 0.020]** |

CPE and 95% confidence intervals from Cox proportional hazards models for incident CAD, PAD, and AIS (n = 4,172). Models include MRS alone, clinical risk factors alone, and clinical risk factors plus MRS. ΔCPE represents the difference in CPE between the clinical + MRS and clinical risk factor models for each ASCVD subtype. CAD, PAD, and AIS groups were not mutually exclusive. The clinical models included body mass index, smoking, HDL-C, LDL-C, systolic blood pressure, lipid-lowering medication use, antihypertensive medication use, and diabetes history. All models were adjusted for age, sex, genetic principal components 1-10, and estimated blood cell proportions. ASCVD, atherosclerotic cardiovascular disease; AIS, acute ischemic stroke; CAD, coronary artery disease; CPE, concordance probability estimate; ΔCPE, change in concordance probability estimate; LRT, likelihood ratio test; MRS, methylation risk score; PAD, peripheral artery disease.
<sup>1</sup>Due to the smaller number of AIS cases, models for AIS were adjusted for a reduced set of base covariates (age, genetic principal components 1-5, and estimated blood cell proportions).
<sup>2</sup>Previously published CAD-specific MRS.<sup>25</sup>

Calibration-in-the-large was similar across clinical risk factor models with and without the inclusion of the MRS. For the 5-year risk, the estimates were 0.0011 for the clinical model, 0.0012 for clinical + MRS-1A, 0.0012 for clinical + MRS-1B, and 0.0011 for clinical + MRS-2. For the 10-year risk, the corresponding estimates were -0.004, -0.004, -0.005, and -0.004, respectively. All values were close to zero, indicating good overall calibration, and the calibration of the clinical risk factor model was not meaningfully altered by the addition of any MRS. Visual inspection of the calibration plots likewise showed good agreement between the predicted and observed risks (**Figure S10**).

We next evaluated risk reclassification following the inclusion of the MRS using categorical NRI (**Table 4**). Across all three MVP-derived MRS, improvements in classification were driven primarily by correct downward reclassification among non-cases, with consistently positive and statistically significant non-event NRI estimates (NRI-) for both the 5-year and 10-year risks. In contrast, event NRI estimates (NRI+) were small and not statistically significant for any model at either time horizon, with confidence intervals spanning zero. The estimates were highly consistent across the 5-and 10-year horizons, as expected, given the use of scaled risk thresholds.

**Table 4.**
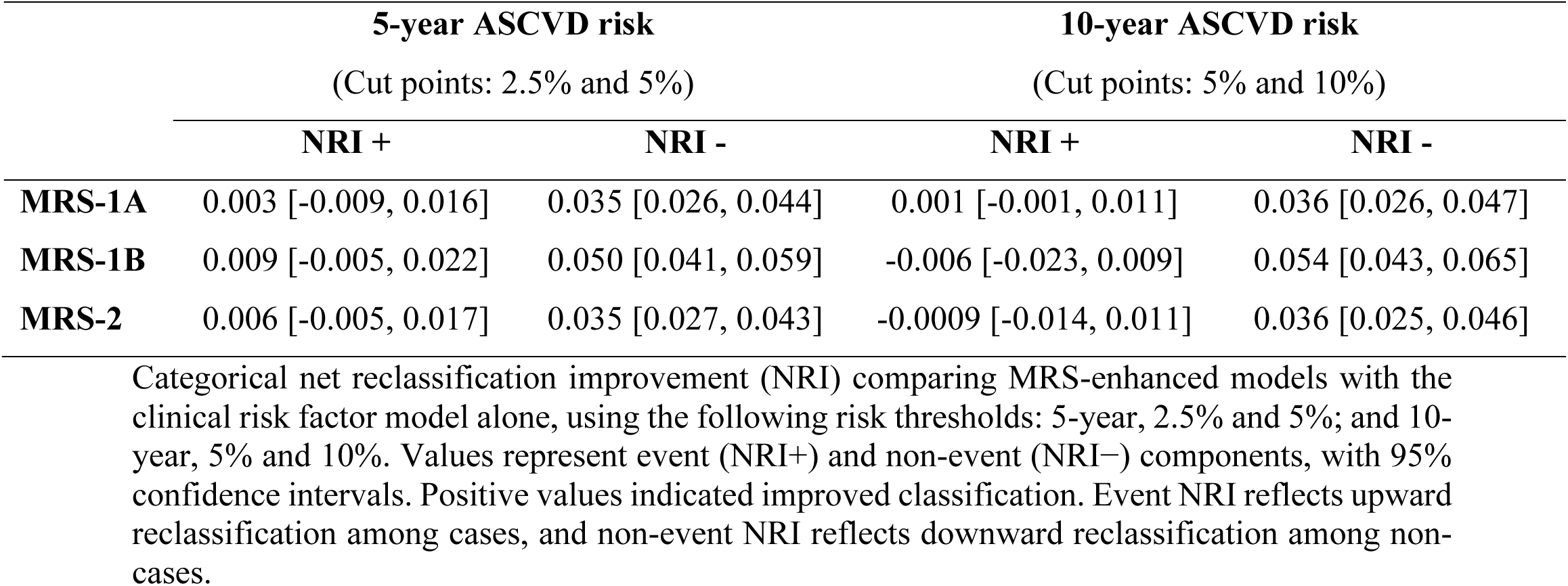
Categorical net reclassification improvement (NRI) comparing MRS-enhanced models with the clinical risk factor model for ASCVD risk.

### ASCVD MRS prediction beyond epigenetic clock GrimAge2

All MRS (MVP and Westerman) were strongly correlated with GrimAge2 (r = 0.81–0.91) (**Figure S11**). Despite these strong correlations, there was little overlap between the CpGs comprising each MRS and GrimAge2 (**Figure S12**). Correlations with the 10 GrimAge2 components, which are DNA methylation-based surrogates for smoking pack-years, C-reactive protein, hemoglobin A1c, cystatin C, tissue inhibitor of metalloproteinases 1, plasminogen activator inhibitor 1, leptin, growth differentiation factor 15, beta-2 microglobulin, and adrenomedullin, varied widely, ranging from 0.11 to 0.86 (**Figure S11**).

As expected, GrimAge2 was a strong predictor of incident ASCVD (HR, 2.43; 95% CI, 2.05-2.89), including after adjustment for clinical risk factors (HR, 2.10; 95% CI, 1.70-2.59). To evaluate whether MRS provided information beyond GrimAge2, we examined models including MRS adjusted for GrimAge2, as well as models further adjusted for clinical risk factors. Across both MVP-derived and CAD-specific MRS, the inclusion of GrimAge2 attenuated MRS effect estimates, with additional attenuation after adjustment for clinical risk factors. Nevertheless, all MRS remained significantly associated with incident ASCVD (**Figure 5**). Consistent with these findings, the addition of each MRS to models including GrimAge2 alone or GrimAge2 plus clinical risk factors significantly improved model fit (all LRT P < 0.05). Similar patterns were observed across ASCVD subtypes, with MVP-derived MRS remaining associated with incident CAD, PAD, and AIS after adjustment for GrimAge2 and clinical risk factors, with attenuated effect estimates consistent with the overall ASCVD findings (**Figure S13**).

**Figure 5.**
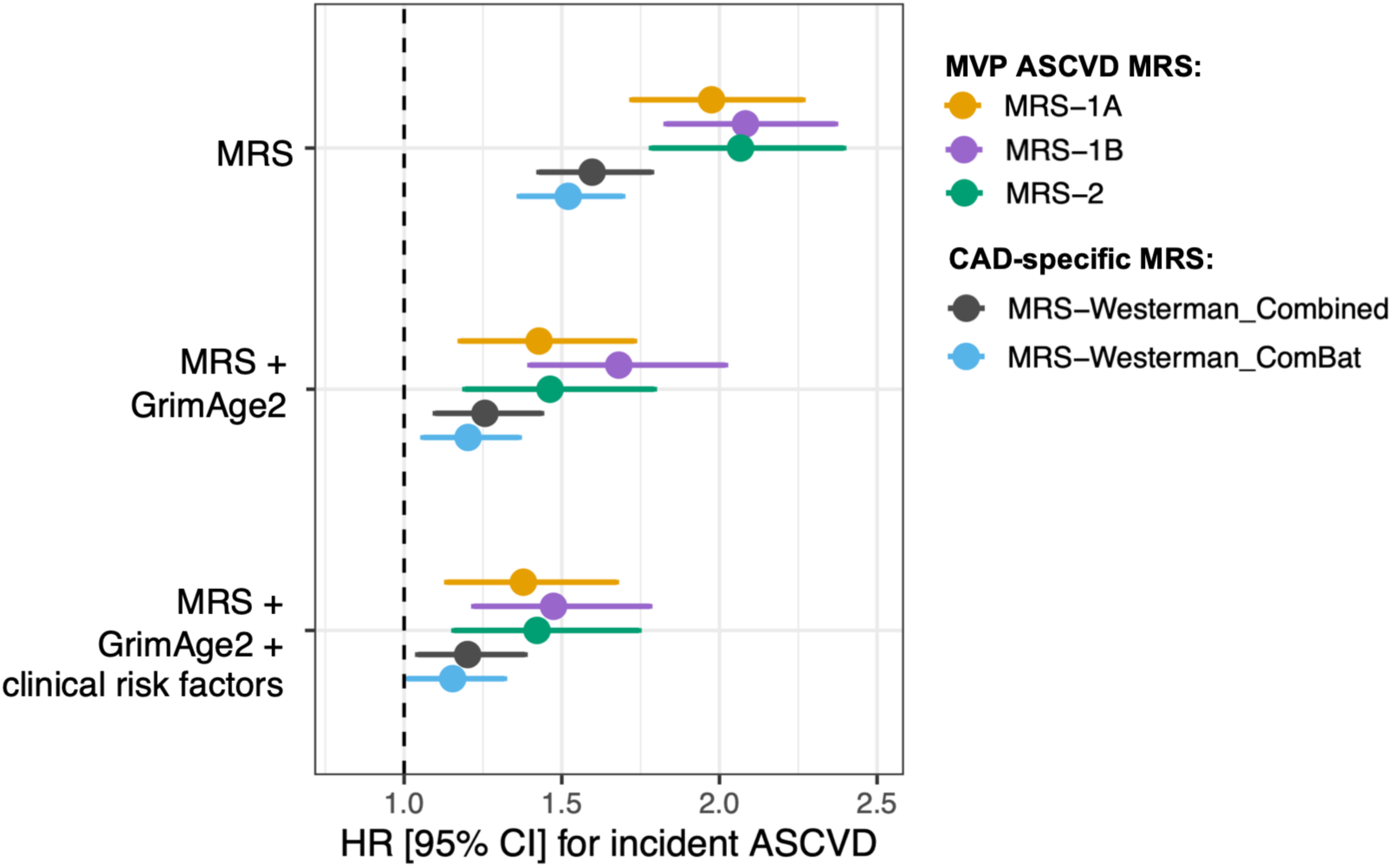
Associations of methylation risk scores (MRS) with incident ASCVD with and without adjustment for GrimAge2 and clinical risk factors. Hazard ratios (HRs) and 95% confidence intervals from Cox proportional hazards models for incident ASCVD are shown per 1-SD increase in each MRS. The clinical risk factors included body mass index, smoking status, HDL-C, LDL-C, systolic blood pressure, lipid-lowering medication use, antihypertensive medication use, and history of diabetes. All models were adjusted for age, sex, genetic principal components 1-10, and estimated blood cell proportions.

## Discussion

In this large prospective study leveraging leukocyte genome-wide DNA methylation data from the MVP, we developed and evaluated three MRS for incident ASCVD and, to our knowledge, provided the first direct evaluation of the incremental predictive value of DNA methylation beyond established clinical risk factors in a primary prevention multi-ancestry cohort. All three MRS were strongly associated with incident ASCVD, modestly improved risk prediction beyond established risk factors while maintaining good model calibration, and demonstrated consistent predictive performance despite being derived using distinct CpG filtering strategies. The comparable performance of the newly developed ASCVD-specific MRS and externally developed CAD-specific MRS supports the robustness of methylation-based predictors of cardiovascular risk in this study.

Prior epigenetic studies have focused on identifying CpGs associated with cardiovascular traits or diseases, with fewer studies evaluating whether methylation-derived signatures improve risk prediction when added to established clinical risk factors,^20–26^ limiting insight into whether these signals capture complementary information. Our findings address this gap by demonstrating that DNA methylation signatures yield modest but consistent incremental predictive value beyond that of traditional risk factors. The magnitude of these improvements is comparable to that observed for other molecular biomarkers, including proteomic and polygenic risk scores, which similarly show gains in discrimination on the order of ∼0.01-0.02 when added to clinical risk factor models.^8,9,11,12^ This pattern likely reflects a ceiling effect, as models incorporating traditional risk factors already explain a substantial proportion of ASCVD risk.^6,7^ Accordingly, the consistent, though modest, gains observed across diverse omics platforms support a broader framework in which multiple, partially independent molecular layers contribute complementary information for risk prediction.

Additional support for the incremental value of these methylation-based signatures was provided by reclassification and calibration analysis. The MRS modestly improved risk stratification, primarily by more accurately identifying individuals at a lower risk, with limited impact on the upward reclassification of those who subsequently developed ASCVD. Non-event NRIs ranged from 3.6% to 5.4%, whereas event NRIs were minimal, a pattern observed in prior proteomics-based ASCVD prediction studies.^12^ Importantly, the inclusion of the MRS did not adversely affect model calibration, indicating good agreement between the predicted and observed risks. Together, these findings further demonstrate that leukocyte DNA methylation signatures capture information complementary to traditional risk factors and support the further evaluation of integrated multi-omics approaches for refining cardiovascular risk prediction.

GrimAge2, a DNA methylation-based predictor of mortality,^42^ was selected for benchmarking because it has demonstrated the strongest associations with cardiovascular disease outcomes compared with other epigenetic clocks.^43–45^ Adjusting for GrimAge2 attenuated the associations of the MRS, indicating that it captures a meaningful portion of the underlying methylation signal. This overlap is expected, as GrimAge2 was constructed using DNA methylation-based surrogates of circulating biomarkers and lifestyle exposures strongly associated with ASCVD risk, including smoking pack-years, cystatin C, high-sensitivity C-reactive protein, and hemoglobin A1c.^42^ Nonetheless, despite this shared signal, all MRS remained independently associated with ASCVD after adjustment for GrimAge2 alone and GrimAge2 plus clinical risk factors, suggesting that they capture additional biological information beyond that reflected by GrimAge2.

The three CpG filtering strategies used to construct the MVP MRS yielded broadly similar predictive performances, making it difficult to clearly favor one approach. The MRS-1B model, derived from CpGs identified in the prevalent ASCVD EWAS without additional reliability-based restrictions, showed slightly stronger associations and modestly improved performance in some analyses, including the MRS-only models and models adjusted for GrimAge2. In contrast, the MRS-1A and MRS-2 models incorporated filtering based on probe reliability,^40^ which may enhance the reproducibility across platforms and studies. Notably, all three MVP-derived models were relatively compact, comprising 49-124 CpGs, suggesting that effective methylation-based predictors of ASCVD can be constructed using relatively small CpG sets. In comparison, the Westerman CAD-MRS scores were constructed using substantially larger CpG sets (300-800 CpGs) but did not demonstrate improved predictive performance; similarly, expanding CpG inclusion beyond 300 sites in our sensitivity analyses did not enhance performance. Taken together, these findings suggest that multiple CpG selection strategies can produce MRS with comparable performance, with potential trade-offs between maximizing predictive performance and prioritizing reproducibility.

This study has several strengths. First, the analyses were conducted in one of the largest cohorts with genome-wide DNA methylation data linked to longitudinal EHR data, enabling the evaluation of incident ASCVD events in a large, well-characterized population with diverse ancestry representation. Second, we used an independent subset for epigenome-wide discovery and model development, thereby reducing the risk of overfitting and enhancing the credibility of the predictive models. Third, we applied multiple complementary CpG filtering strategies and penalized regression methods to construct parsimonious MRS while maintaining robust predictive performance. Finally, a comparison with a previously published CAD-specific MRS^25^ and a well-validated epigenetic aging biomarker^42^ provided important context for interpreting the incremental value of ASCVD-specific methylation signatures.

Key limitations should also be considered in this study. First, DNA methylation was measured in peripheral blood samples, which may not fully capture disease-relevant epigenetic changes in vascular tissues. Second, the MVP cohort primarily consisted of older male U.S. Veterans, which may limit the generalizability of the results to other populations, particularly females and younger individuals. The limited number of incident ASCVD events among females precluded adequately powered sex-stratified analysis. Third, although internal validation was performed using independent training and testing subsets, and the externally developed CAD-specific MRS demonstrated similar performance in a head-to-head comparison, external validation of the ASCVD-specific MRS is important to confirm their generalizability. Finally, although the prospective design strengthens the interpretation, DNA methylation remains a dynamic biomarker that may influence and reflect the underlying disease processes.

In summary, DNA methylation signatures were strongly associated with incident ASCVD and provided modest improvements in risk prediction beyond established clinical risk factors in a large prospective cohort study. Much of the predictive signal captured by these MRS overlapped with the epigenetic aging biomarker GrimAge2, although all scores retained independent associations with ASCVD. More broadly, these findings highlight the potential value of integrating DNA methylation measurements into future multi-omics frameworks for cardiovascular disease risk prediction and targeted prevention.

## Acknowledgements

We gratefully acknowledge the Veterans who participated in the VA’s Million Veteran Program and the contributions of the Million Veteran Program Core members listed in **Text S1.**

## Data Availability

Access to MVP data is governed by the scope of the MVP informed consent and VA policies. MVP data access requires a scientific review by the appropriate VA review committees. More information on applying for access to MVP data is available at: https://www.research.va.gov/MVP/research.cfm.

## Code Availability

The code and weights used to calculate the methylation risk scores developed in this study are available at https://github.com/alexabarad/MVP_ASCVD_MRS.

## Abbreviations

AFR: African ancestry
AIS: acute ischemic stroke
AMR: Admixed American ancestry
ASCVD: atherosclerotic cardiovascular disease
CAD: coronary artery disease
CKD: chronic kidney disease
CpG: cytosine-phosphate-guanine dinucleotides
EAS: East Asian ancestry
EHR: electronic health record
EUR: European ancestry
EWAS: epigenome-wide association study
HDL-C: high-density lipoprotein cholesterol
KEGG: Kyoto Encyclopedia of Genes and Genomes
LDL-C: low-density lipoprotein cholesterol
LRT: likelihood ratio test
MRS: methylation risk score
MVP: Million Veteran Program
PAD: peripheral artery disease
PTSD: post-traumatic stress disorder
SAS: South Asian ancestry
VA: Veterans Affairs
VHA: Veterans Health Administration.

