## Supplementary Material 1 for "Leukocyte DNA methylation-based signatures for atherosclerotic cardiovascular disease risk prediction in the Million Veteran Program"

**Supplementary Material 1:** Supplemental Tables S1-S6, Supplemental Figures S1-S13, Supplemental Text S1.

**Supplementary Material 2:** Annotated genome-wide significant probes from prevalent and incident ASCVD EWAS.

**Supplementary Material 3:** Elastic-net coefficients used to compute each methylation risk score.

**Table S1. Comparison of descriptive characteristics of the pseudorandom methylation incident cohort and the genotyped MVP (R4) after exclusion of prevalent ASCVD.**

| | **Full genotyped MVP (R4)** (n=462864) | | **Incident methylation sample (“pseudorandom” sample)** (n=17114) | |
|---|---|---|---|---|
| | Value | n missing | Value | n missing |
| Age, y | 58.7 ± 14.4 | 2 | 58.9 ± 14.1 | 0 |
| Males, n (%) | 410404 (88.7) | 0 | 15342 (89.6) | 0 |
| Genetically inferred ancestry, n (%) | | 0 | | 0 |
| African | 93865 (20.3) | | 5067 (29.6) | |
| Admixed American | 48671 (10.5) | | 2129 (12.4) | |
| East Asian | 5955 (1.3) | | 307 (1.8) | |
| European | 306973 (66.3) | | 9272 (54.2) | |
| South Asian | 502 (0.1) | | 22 (0.1) | |
| Unidentified | 6898 (1.5) | | 317 (1.9) | |
| Body mass index, kg/m$^2$ | 29.7 ± 5.8 | 3492 | 29.7 ± 5.8 | 113 |
| Smoking status, n (%) | | 25528 | | 1135 |
| Current | 118991 (27.2) | | 4640 (29.0) | |
| Former | 184728 (42.2) | | 6705 (42.0) | |
| Never | 133617 (30.6) | | 4634 (29.0) | |
| History of hypertension, n (%) | 190440 (41.2) | 97 | 6829 (39.9) | 0 |
| History of diabetes, n (%) | 96846 (20.9) | 0 | 3603 (21.1) | 0 |
| History of CKD, n (%) | 28798 (6.2) | 97 | 1052 (6.1) | 0 |
| History of PTSD, n (%) | 76557 (16.5) | 97 | 2851 (16.7) | 0 |
| Diabetes medication use, n (%) | 80725 (17.4) | 0 | 3035 (17.7) | 0 |
| Lipid lowering medication use, n (%) | 175598 (37.9) | 0 | 6584 (38.5) | 0 |
| Hypertension medication use, n (%) | 251156 (54.3) | 0 | 9582 (56.0) | 0 |
| LDL-C, mg/dL | 105 ± 34.5 | 63691 | 105 ± 34.2 | 2502 |
| HDL-C, mg/dL | 47.6 ± 14.9 | 63691 | 47.7 ± 14.8 | 2502 |
| SBP, mm Hg | 131 ± 16.2 | 13917 | 131 ± 16.2 | 556 |
| Incident ASCVD, n (%) | 73940 (16.0) | 0 | 2789 (16.3) | 0 |
| CAD | 44486 (9.6) | | 1703 (9.9) | |
| PAD | 23901 (5.2) | | 867 (5.1) | |
| AIS | 5571 (1.2) | | 219 (1.3) | |
| Incident CAD, n (%) | 49690 (10.7) | 0 | 1916 (11.2) | 0 |
| Incident PAD, n (%) | 28331 (6.1) | 0 | 1033 (6.0) | 0 |
| Incident AIS, n (%) | 6613 (1.4) | 0 | 254 (1.5) | 0 |
| >1 incident ASCVD subtype, n (%) | 10225 (2.2) | 0 | 398 (2.3) | 0 |

MVP genotyped (R4) subset includes participants enrolled before 2020 to match the enrollment period of the methylation subset and excludes individuals with prevalent ASCVD. AIS, acute ischemic stroke; CAD, coronary artery disease; CKD, chronic kidney disease; HDL-C, high-density lipoprotein cholesterol; LDL-C, low-density lipoprotein cholesterol; PAD, peripheral artery disease; PTSD, post-traumatic stress disorder; SBP, systolic blood pressure.

**Table S2. Incident cohort descriptive characteristics.**

| | All (n = 17114) | | Training set (n = 11980) | | Testing set (n = 5134) | |
|---|---|---|---|---|---|---|
| | Value | n missing | Value | n missing | Value | n missing |
| Age, y | 58.9 ± 14.1 | 0 | 58.8 ± 14.1 | 0 | 59.2 ± 14.1 | 0 |
| Males, n (%) | 15342 (89.6) | 0 | 10746 (89.7) | 0 | 4596 (89.5) | 0 |
| Genetically inferred ancestry, n (%) | | 0 | | 0 | | 0 |
| African | 5067 (29.6) | | 3544 (29.6) | | 1523 (29.7) | |
| Admixed American | 2129 (12.4) | | 1486 (12.4) | | 643 (12.5) | |
| East Asian | 307 (1.8) | | 213 (1.8) | | 94 (1.8) | |
| European | 9272 (54.2) | | 6514 (54.4) | | 2758 (53.7) | |
| South Asian | 22 (0.1) | | 19 (0.2) | | 3 (0.1) | |
| Unidentified | 317 (1.9) | | 204 (1.7) | | 113 (2.2) | |
| Body mass index, $kg/m^2$ | 29.7 ± 5.8 | 113 | 29.7 ± 5.8 | 76 | 29.8 ± 5.8 | 37 |
| Smoking status, n (%) | | 1135 | | 785 | | 350 |
| Current | 4640 (29.0) | | 3249 (29.0) | | 1391 (29.1) | |
| Former | 6705 (42.0) | | 4685 (41.8) | | 2020 (42.2) | |
| Never | 4634 (29.0) | | 3261 (29.1) | | 1373 (28.7) | |
| History of hypertension, n (%) | 6829 (39.9) | 0 | 4805 (40.1) | 0 | 2024 (39.4) | 0 |
| History of diabetes, n (%) | 3603 (21.1) | 0 | 2523 (21.1) | 0 | 1080 (21.0) | 0 |
| History of CKD, n (%) | 1052 (6.1) | 0 | 739 (6.2) | 0 | 313 (6.1) | 0 |
| History of PTSD, n (%) | 2851 (16.7) | 0 | 1977 (16.5) | 0 | 874 (17.0) | 0 |
| Diabetes medication use, n (%) | 3035 (17.7) | 0 | 2114 (17.6) | 0 | 921 (17.9) | 0 |
| Lipid lowering medication use, n (%) | 6584 (38.5) | 0 | 4616 (38.5) | 0 | 1968 (38.3) | 0 |
| Hypertension medication use, n (%) | 9582 (56.0) | 0 | 6720 (56.1) | 0 | 2862 (55.7) | 0 |
| LDL-C, mg/dL | 105 ± 34.2 | 2502 | 105 ± 34.1 | 1723 | 106 ± 34.3 | 779 |
| HDL-C, mg/dL | 47.7 ± 14.8 | 2502 | 47.7 ± 14.8 | 1723 | 47.8 ± 14.8 | 779 |
| SBP, mm Hg | 131 ± 16.2 | 556 | 131 ± 16.3 | 373 | 130 ± 15.9 | 183 |
| Time-to-ASCVD, y | 7.4 [4.6, 9.5] | 203 | 7.4 [4.6, 9.6] | 133 | 7.4 [4.6, 9.5] | 70 |
| Incident ASCVD, n (%) | 2789 (16.3) | 0 | 1955 (16.3) | 0 | 834 (16.3) | 0 |
| CAD | 1703 (9.9) | | 1197 (10.0) | | 506 (9.9) | |
| PAD | 867 (5.1) | | 601 (5.0) | | 266 (5.2) | |
| AIS | 219 (1.3) | | 157 (1.3) | | 62 (1.2) | |
| Incident CAD, n (%) | 1916 (11.2) | 0 | 1344 (11.2) | 0 | 572 (11.1) | 0 |
| Incident PAD, n (%) | 1033 (6.0) | 0 | 718 (6.0) | 0 | 315 (6.1) | 0 |
| Incident AIS, n (%) | 254 (1.5) | 0 | 182 (1.5) | 0 | 72 (1.4) | 0 |
| >1 incident ASCVD subtype, n (%) | 398 (2.3) | 0 | 275 (2.3) | 0 | 123 (2.4) | 0 |

AIS, acute ischemic stroke; CAD, coronary artery disease; CKD, chronic kidney disease; HDL-C, high-density lipoprotein cholesterol; LDL-C, low-density lipoprotein cholesterol; PAD, peripheral artery disease; PTSD, post-traumatic stress disorder; SBP, systolic blood pressure.

**Table S3. Prevalent ASCVD cohort characteristics stratified by cases and controls.**

| | **Prevalent ASCVD cases** (n=13780) | | **Controls** (n=13780) | |
|---|---|---|---|---|
| | Value | n missing | Value | n missing |
| Age, y | 71.4 ± 10.6 | 0 | 64.6 ± 14.6 | 0 |
| Males, n (%) | 13398 (97.2) | 0 | 12696 (92.1) | 0 |
| Genetically inferred ancestry, n (%) | | 0 | | 0 |
| African | 2294 (21.7) | | 3568 (25.9) | |
| Admixed American | 1031 (7.5) | | 1611 (11.7) | |
| East Asian | 95 (0.7) | | 190 (1.4) | |
| European | 9507 (69.0) | | 8184 (59.4) | |
| South Asian | 5 (0.04) | | 15 (0.1) | |
| Unidentified | 148 (1.1) | | 212 (1.5) | |
| BMI, kg/m$^2$ | 29.5 (5.9) | 31 | 29.5 (5.8) | 90 |
| Smoking status, n (%) | | 269 | | 829 |
| Current | 3312 (24.5) | | 3279 (25.3) | |
| Former | 7195 (53.3) | | 6012 (46.4) | |
| Never | 3004 (22.2) | | 3660 (28.3) | |
| History of hypertension, n (%) | 7051 (51.2) | 5 | 5982 (43.4) | 9 |
| History of diabetes, n (%) | 5895 (42.8) | 0 | 4224 (30.7) | 0 |
| History of CKD, n (%) | 2099 (15.2) | 5 | 1106 (15.2) | 9 |
| History of PTSD, n (%) | 1551 (11.3) | 5 | 2171 (15.8) | 9 |
| Diabetes medication use, n (%) | 5078 (36.9) | 0 | 3534 (25.6) | 0 |
| Lipid lowering medication use, n (%) | 10308 (74.8) | 0 | 6162 (44.7) | 0 |
| Hypertension medication use, n (%) | 11441 (83.0) | 0 | 5982 (43.4) | 0 |
| Prevalent CAD, n (%) | 9952 (72.2) | 0 | 0 | 0 |
| Prevalent PAD, n (%) | 8175 (59.3) | 0 | 0 | 0 |
| Prevalent AIS, n (%) | 2497 (18.1) | 0 | 0 | 0 |
| >1 prevalent ASCVD subtype, n (%) | 3792 (27.5) | 0 | 0 | 0 |
| Age at first reported diagnosis, y | 64.7 ± 10.5 | 824 | - | - |
| Time from diagnosis to blood draw, y | 6.6 [3.0, 10.2] | 824 | - | - |

AIS, acute ischemic stroke; CAD, coronary artery disease; CKD, chronic kidney disease; HDL-C, high-density lipoprotein cholesterol; LDL-C, low-density lipoprotein cholesterol; PAD, peripheral artery disease; PTSD, post-traumatic stress disorder; SBP, systolic blood pressure.

**Table S4. Significantly enriched KEGG pathways among genes annotated to Bonferroni-significant CpG sites identified in the EWAS of prevalent ASCVD.**

| KEGG Pathway | Pathway description | Genes in pathway | Input genes in pathway | FDR |
|---|---|---|---|---|
| hsa04668 | TNF signaling pathway | 114 | 31 | 4.48E-02 |
| hsa04062 | Chemokine signaling pathway | 183 | 47 | 4.48E-02 |
| hsa04210 | Apoptosis | 132 | 35 | 4.48E-02 |
| hsa05223 | Non-small cell lung cancer | 71 | 26 | 4.48E-02 |
| hsa05203 | Viral carcinogenesis | 184 | 45 | 4.48E-02 |

Kyoto Encyclopedia of Genes and Genomes (KEGG) pathway enrichment analysis was performed using the *gometh* function in the missMethyl R package, which accounts for the unequal representation of genes on the Illumina EPIC methylation array. The analysis included 5,403 Bonferroni-significant CpG sites identified in the prevalent ASCVD EWAS, using all 721,647 CpG sites that passed quality control as the background. FDR-adjusted P values were calculated using the Benjamini-Hochberg method. "Genes in pathway" denotes the total number of genes annotated to each KEGG pathway, and "Input genes in pathway" denotes the number of genes annotated to the Bonferroni-significant CpG sites. Only pathways with FDR < 0.05 are shown.

**Table S5. Sensitivity analyses of penalized regression specifications for methylation risk score (MRS) construction and corresponding AUCs for incident ASCVD prediction.**

| Approach | Penalized regression settings | N CpGs | MRS | Clinical | Clinical+ MRS |
|---|---|---|---|---|---|
| 1A | α= 0.5 λ_1SE | 49 | 0.708 (0.691, 0.724) | 0.709 (0.690, 0.727) | 0.721 (0.703, 0.738) |
| | α= 0.5 λ_min | 237 | 0.713 (0.697, 0.730) | 0.709 (0.690, 0.727) | 0.716 (0.698, 0.734) |
| | Flexible α λ_1SE | 130 (α = 0.7) | 0.715 (0.699, 0.732) | 0.709 (0.690, 0.727) | 0.722 (0.704, 0.739) |
| 1B | α= 0.5 λ_1SE | 124 | 0.717 (0.701, 0.733) | 0.709 (0.690, 0.727) | 0.722 (0.704, 0.740) |
| | α= 0.5 λ_min | 360 | 0.715 (0.699, 0.732) | 0.709 (0.690, 0.727) | 0.712 (0.693, 0.730) |
| | Flexible α λ_1SE | 354 (α = 0.1) | 0.721 (0.705, 0.738) | 0.709 (0.690, 0.727) | 0.721 (0.703, 0.740) |
| 2 | α= 0.5 λ_1SE | 49 | 0.705 (0.688, 0.721) | 0.709 (0.690, 0.727) | 0.720 (0.702, 0.738) |
| | α= 0.5 λ_min | 212 | 0.711 (0.694, 0.727) | 0.709 (0.690, 0.727) | 0.714 (0.696, 0.732) |
| | Flexible α λ_1SE | 138 (α = 0.1) | 0.706 (0.690, 0.723) | 0.709 (0.690, 0.727) | 0.719 (0.702, 0.737) |

Area under the curve (AUC) values and 95% confidence intervals from logistic regression models comparing MRS-only models, clinical risk factor models, and combined clinical + MRS models across alternative elastic net specifications. Sensitivity analyses evaluated different alpha (α = 0.1-1) and lambda (λ; λ_1SE and λ_min) parameters used for penalized regression in the construction of the MRS. Clinical models include body mass index, smoking, HDL-C, LDL-C, systolic blood pressure, lipid-lowering medication use, antihypertensive medication use, and diabetes history. All models were adjusted for age, sex, genetic principal components 1-10, and estimated blood cell proportions. λ_min denotes the value of λ corresponding to the minimum cross-validation error. λ_1SE denotes the largest value of λ within one standard error of the minimum cross-validation error.

**Table S6. Incremental predictive value of methylation risk scores (MRS) for incident ASCVD and its subtypes, excluding deaths (sensitivity analyses).**

| | | ASCVD (n cases= 651) | CAD (n cases = 414) | PAD (n cases = 285) | AIS (n cases = 59)[1] |
|---|---|---|---|---|---|
| **MRS** | **Model** | **CPE [95% CI]** | **CPE [95% CI]** | **CPE [95% CI]** | **CPE [95% CI]** |
| MVP 1a | **MRS** | 0.687 [0.669, 0.706] | 0.658 [0.634, 0.682] | 0.730 [0.706, 0.754] | 0.722 [0.663, 0.781] |
| | **Clinical** | 0.700 [0.681, 0.718] | 0.688 [0.664, 0.713] | 0.738 [0.714, 0.762] | 0.718 [0.655, 0.782] |
| | **Clinical + MRS** | 0.714 [0.696, 0.732] | 0.694 [0.670, 0.717] | 0.756 [0.733, 0.779] | 0.744 [0.684, 0.804] |
| | **ΔCPE [95% CI]** | **0.014 [0.007, 0.022]** | **0.006 [0.0006, 0.015]** | **0.018 [0.007, 0.030]** | **0.026 [0.003, 0.048]** |
| MVP 1b | **MRS** | 0.694 [0.676, 0.713] | 0.667 [0.644, 0.691] | 0.736 [0.712, 0.762] | 0.726 [0.665, 0.786] |
| | **Clinical** | 0.700 [0.681, 0.718] | 0.688 [0.664, 0.713] | 0.738 [0.714, 0.762] | 0.718 [0.655, 0.782]] |
| | **Clinical + MRS** | 0.716 [0.698, 0.734] | 0.697 [0.674, 0.721] | 0.758 [0.735, 0.781] | 0.744 [0.683, 0.805] |
| | **ΔCPE [95% CI]** | **0.016 [0.007, 0.025]** | **0.009 [0.002, 0.019]** | **0.020 [0.007, 0.032]** | **0.026 [0.002, 0.050]** |
| MVP 2 | **MRS** | 0.683 [0.664, 0.702] | 0.654 [0.630, 0.678] | 0.726 [0.702, 0.751] | 0.725 [0.666, 0.783] |
| | **Clinical** | 0.700 [0.681, 0.718] | 0.688 [0.664, 0.713] | 0.738 [0.714, 0.762] | 0.718 [0.655, 0.782]] |
| | **Clinical + MRS** | 0.713 [0.695, 0.730] | 0.692 [0.669, 0.716] | 0.756 [0.733, 0.779] | 0.747 [0.688, 0.806] |
| | **ΔCPE [95% CI]** | **0.013 [0.005, 0.020]** | **0.004 [0.0001, 0.014]** | **0.018 [0.006, 0.029]** | **0.029 [0.003, 0.053]** |
| Westerman MRS-Combined[2] | **MRS** | 0.681 [0.662, 0.699] | 0.654 [0.631, 0.678] | 0.720 [0.695, 0.744] | 0.684 [0.622, 0.747] |
| | **Clinical** | 0.700 [0.681, 0.718] | 0.688 [0.664, 0.713] | 0.738 [0.714, 0.762] | 0.718 [0.655, 0.782] |
| | **Clinical + MRS** | 0.710 [0.693, 0.728] | 0.691 [0.668, 0.715] | 0.753 [0.731, 0.776] | 0.729 [0.668, 0.790] |
| | **ΔCPE [95% CI]** | **0.010 [0.004, 0.017]** | **0.003 [-0.0005, 0.010]** | **0.016 [0.005, 0.025]** | **0.011 [-0.0009, 0.034]** |
| Westerman MRS-ComBat[2] | **MRS** | 0.675 [0.657, 0.694] | 0.650 [0.626, 0.674] | 0.717 [0.691, 0.742] | 0.682 [0.620, 0.744] |
| | **Clinical** | 0.700 [0.681, 0.718] | 0.688 [0.664, 0.713] | 0.738 [0.714, 0.762] | 0.718 [0.655, 0.782] |
| | **Clinical + MRS** | 0.707 [0.689, 0.725] | 0.690 [0.666, 0.714] | 0.751 [0.728, 0.774] | 0.727 [0.667, 0.787] |
| | **ΔCPE [95% CI]** | **0.008 [0.002, 0.014]** | **0.002 [-0.001, 0.009]** | **0.013 [0.003, 0.022]** | **0.009 [-0.016, 0.031]** |

CPE and 95% confidence intervals from Cox proportional hazards models for incident ASCVD, CAD, PAD, AIS, with cases excluding deaths (n total = 4,172). Models include MRS alone, clinical risk factors alone, and clinical risk factors + MRS. ΔCPE represents the difference in CPE between the clinical + MRS model and the clinical risk factor model for each ASCVD subtype. CAD, PAD, AIS groups are not mutually exclusive. Clinical models include body mass index, smoking, HDL-C, LDL-C, systolic blood pressure, lipid-lowering medication use, antihypertensive medication use, and diabetes history. All models were adjusted for age, sex, genetic principal components 1-10, and estimated blood cell proportions. ASCVD, atherosclerotic cardiovascular disease; AIS, acute ischemic stroke; CAD, coronary artery disease; CPE, concordance probability estimate; ΔCPE, change in concordance probability estimate; LRT, likelihood ratio test; MRS, methylation risk score; PAD, peripheral artery disease.

[1]Due to the smaller number of AIS cases, models for AIS were adjusted for a reduced set of base covariates (age, genetic principal components 1-5, and estimated blood cell proportions).

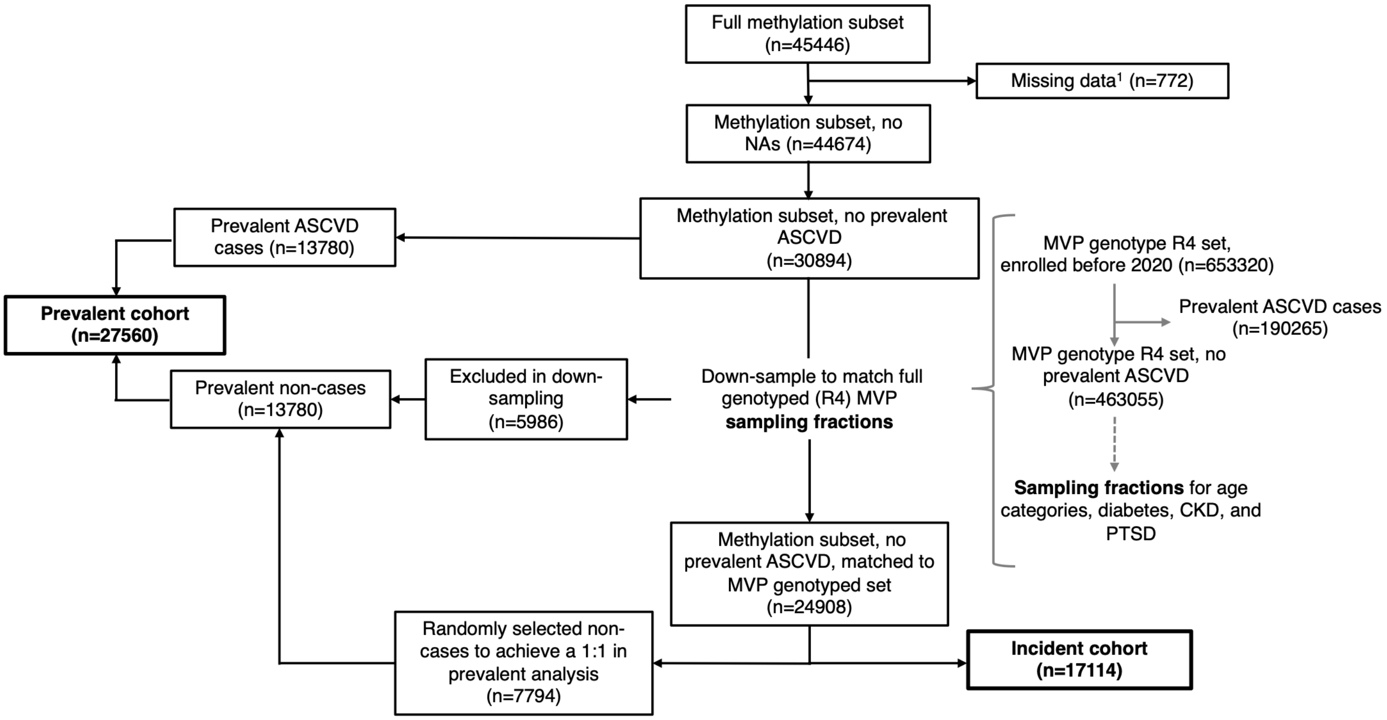


**Figure S1. Flowchart illustrating the generation of the independent incident and prevalent cohorts.** Generation of the pseudorandom cohort was performed using sampling fractions from MVP genotype R4 set, with random downsampling without replacement implemented using the *sample_n* R function. Prevalent ASCVD was defined as a composite of CAD, PAD, or AIS, with a first recorded diagnosis at or before the date of blood collection. [1]Participants with missing data were defined as those missing CAD, PAD, or AIS information, variables required for the resampling procedures (age, diabetes, CKD, or PTSD), or technical covariates (technical PCs, DNA methylation scanner ID, or sample storage duration). AIS, acute ischemic stroke; ASCVD, atherosclerotic cardiovascular disease; CAD, coronary artery disease; CKD, chronic kidney disease; PAD, peripheral artery disease; PTSD, post-traumatic stress disorder.

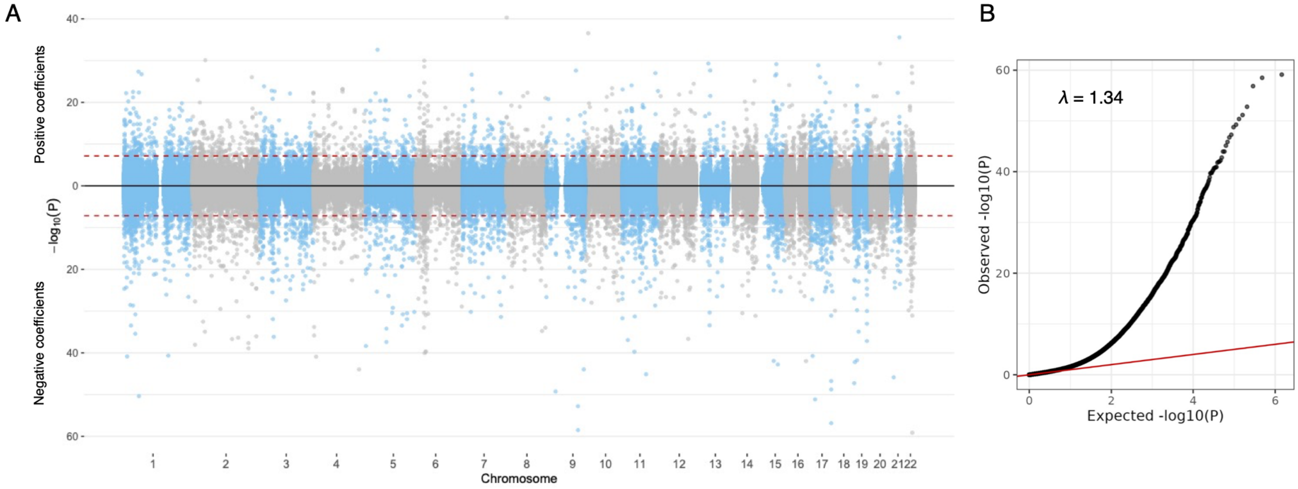


**Figure S2. Miami (A) and quantile–quantile (B) plots for the prevalent ASCVD EWAS.** (A) Positive associations are shown in the upper panel and negative associations in the lower panel. The dashed red line indicates the Bonferroni significance threshold ($6.9x10^{-8}$). (B) Quantile–quantile plot showing genomic inflation (λ) after *bacon* adjustment.

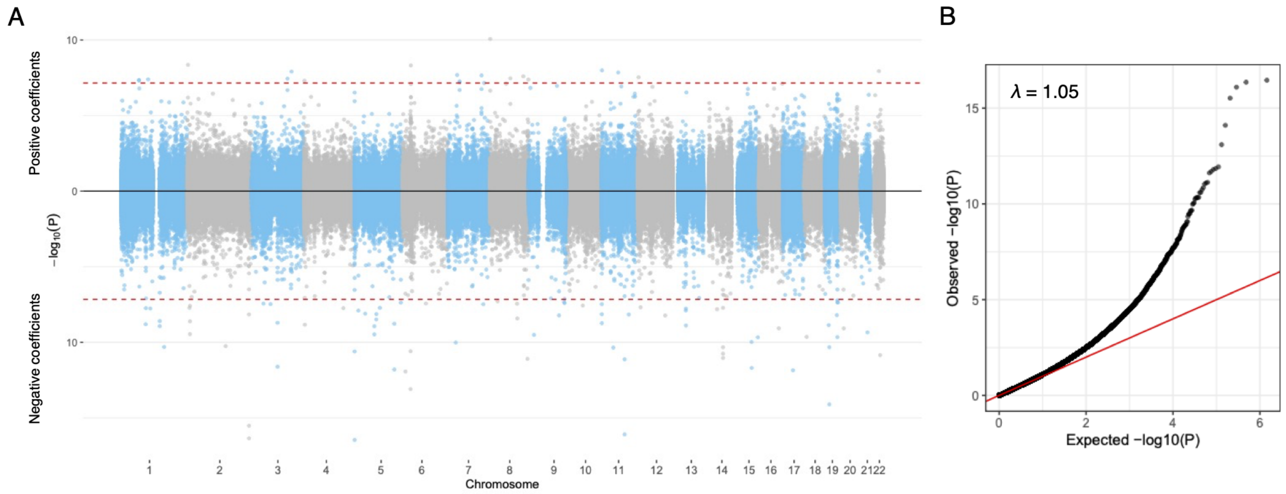


**Figure S3. Miami (A) and quantile–quantile (B) plots for the incident ASCVD EWAS.** (A) Positive associations are shown in the upper panel and negative associations in the lower panel. The dashed red line indicates the Bonferroni significance threshold ($6.9x10^{-8}$). (B) Quantile–quantile plot showing genomic inflation (λ) after *bacon* adjustment.

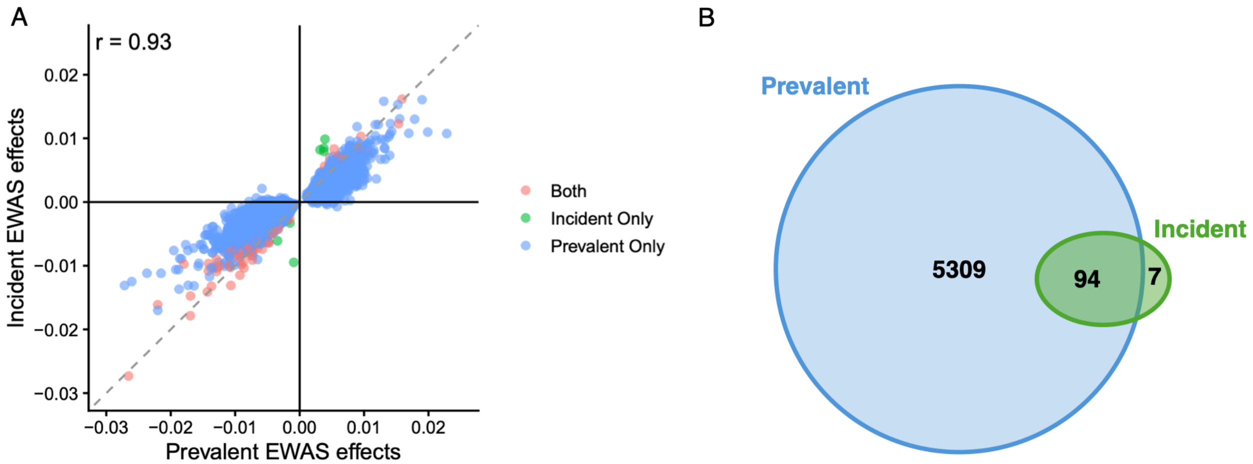


**Figure S4. Agreement between prevalent and incident ASCVD EWAS results.** (A) Pearson correlation between CpG effect estimates from the incident and prevalent EWAS. Only CpGs surpassing the Bonferroni significance threshold in at least one EWAS were included. (B) Venn diagram showing the overlap of Bonferroni-significant CpG sites between analyses. ASCVD, atherosclerotic cardiovascular disease.

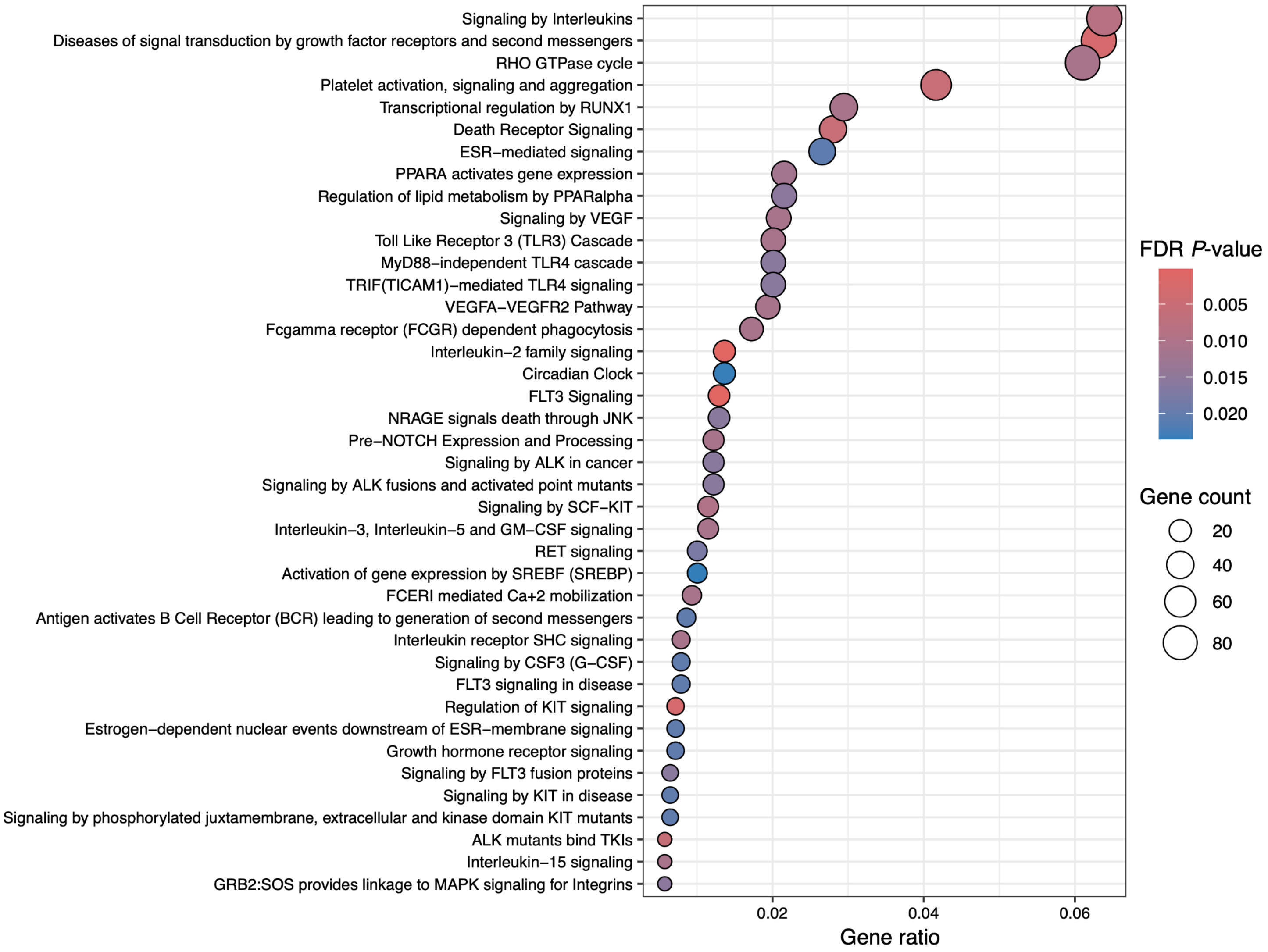


**Figure S5. Top enriched Reactome pathways among genes annotated to Bonferroni-significant CpG sites from the prevalent ASCVD EWAS.** Reactome pathway enrichment analysis was performed using genes annotated to the 5,403 Bonferroni-significant CpG sites identified in the prevalent ASCVD EWAS. The top 40 pathways with a false discovery rate (FDR)-adjusted P value < 0.05 are shown. Dot size represents the number of input genes mapped to each pathway, dot color indicates the FDR-adjusted P value, and the x-axis represents the gene ratio (number of input genes annotated to a pathway divided by the total number of input genes).

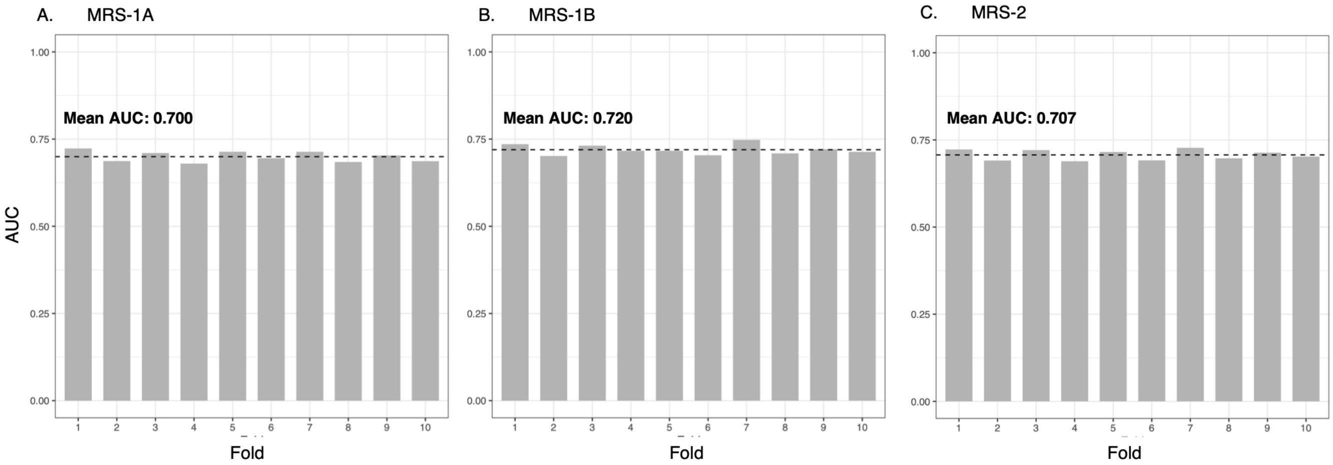


**Figure S6. 10-fold cross-validated discrimination across elastic net approaches.** Results from elastic net models used to construct methylation risk scores (MRS) under three modeling strategies: Approach 1A (A), Approach 1B (B), and Approach 2 (C). Panels show the AUC values across the 10 cross-validation folds, with mean AUC indicated for each approach.

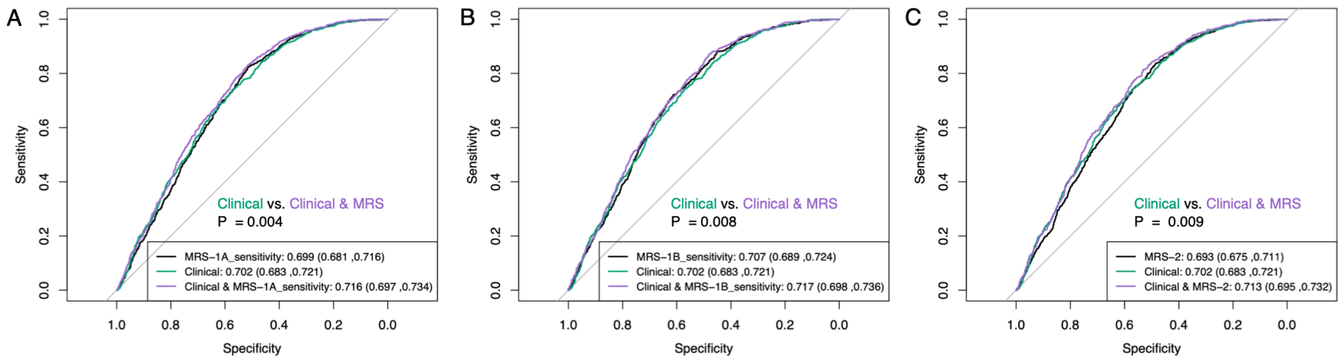


**Figure S7. Discrimination of elastic net-derived methylation risk scores (MRS) for ASCVD excluding deaths (sensitivity analyses).** Receiver operating characteristic curves from logistic regression models comparing MRS-only models, clinical risk factors alone, and combined clinical + MRS models across three elastic net approaches (Panels A–C). AUC values (95% CI) are shown for each model. P-values reflect comparisons of clinical versus clinical + MRS models using DeLong's test. Analyses were conducted in the held-out test set (n = 5,134 with 743 incident ASCVD cases [excluding deaths] for MRS-only models; n = 4,172 with 651 cases [excluding deaths] for clinical and combined models). Clinical models included body mass index, smoking status, HDL-C, LDL-C, systolic blood pressure, lipid-lowering medication use, antihypertensive medication use, and history of diabetes. All models were adjusted for age, sex, genetic principal components 1–10, and estimated blood cell proportions.

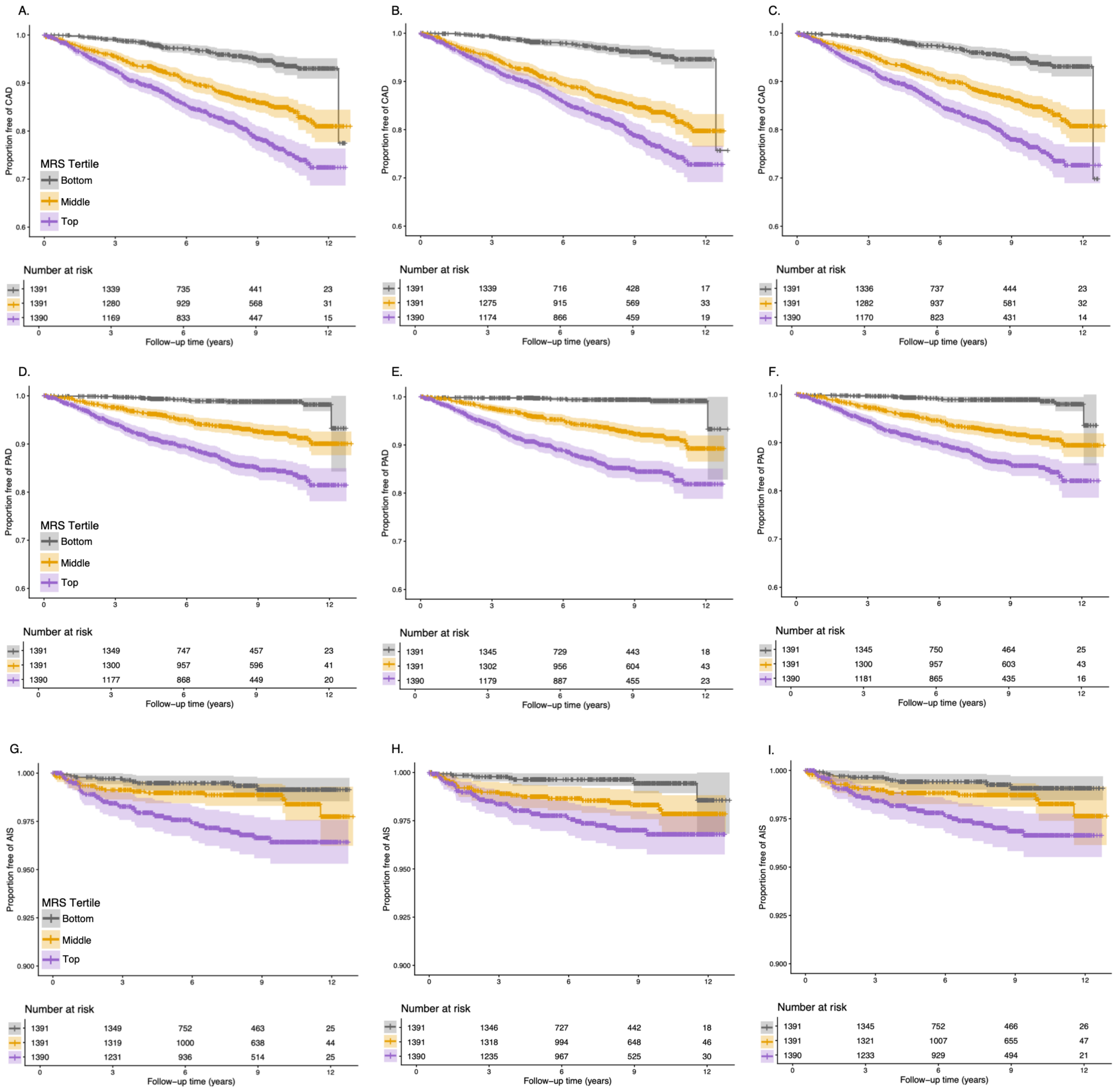


**Figure S8. Kaplan–Meier curves for CAD, PAD, and AIS stratified by methylation risk score (MRS) tertiles.** Kaplan–Meier estimates of disease-free survival across tertiles of the MRS derived from three elastic net approaches. Shaded areas represent 95% confidence intervals. The number of cases was 489 for coronary artery disease (CAD; Panels A–C), 285 for peripheral artery disease (PAD; Panels D–F), and 67 for acute ischemic stroke (AIS; Panels G–I).

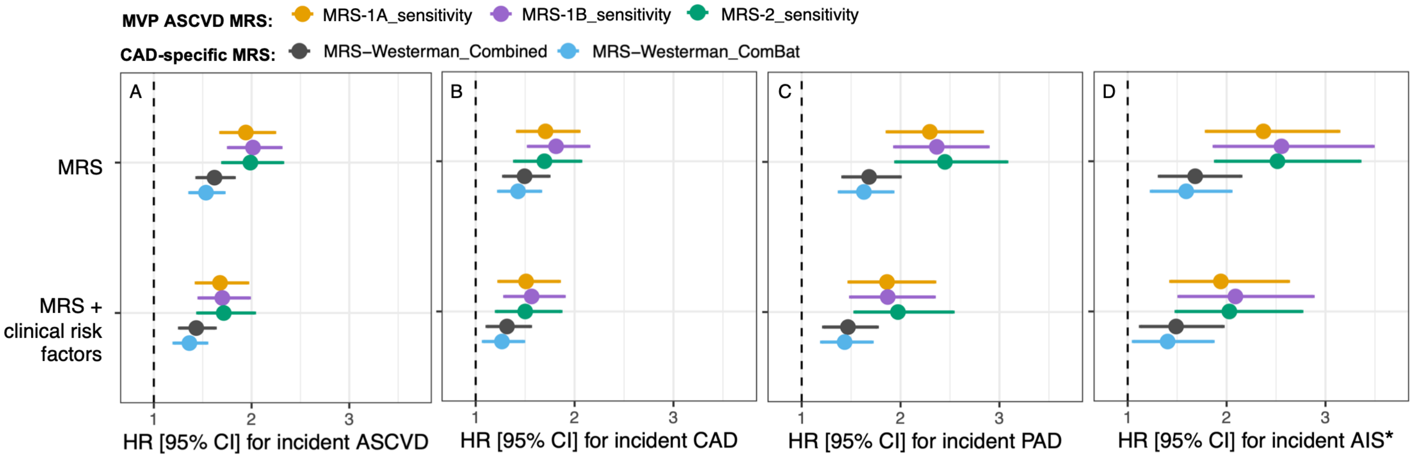


**Figure S9. Associations of methylation risk scores (MRS) with incident ASCVD and its subtypes, excluding deaths (sensitivity analyses).** Hazard ratios (HRs) and 95% confidence intervals from Cox proportional hazards models for incident ASCVD (A), coronary artery disease (CAD) (B), peripheral artery disease (PAD) (C), and acute ischemic stroke (AIS) (D). The number of cases was 651 for ASCVD, 414 for CAD, 285 for PAD, and 59 for AIS. HR [95%CI] are shown per 1-SD increase in each MRS. The top row shows models including the MRS alone, and the bottom row shows models additionally adjusted for clinical risk factors, including body mass index, smoking status, HDL-C, LDL-C, systolic blood pressure, lipid-lowering medication use, antihypertensive medication use, and history of diabetes. All models were adjusted for age, sex, genetic principal components 1-10, and estimated blood cell proportions.

*Due to the smaller number of AIS cases, models for AIS were adjusted for a reduced set of base covariates (age, genetic principal components 1-5, and estimated blood cell proportions).

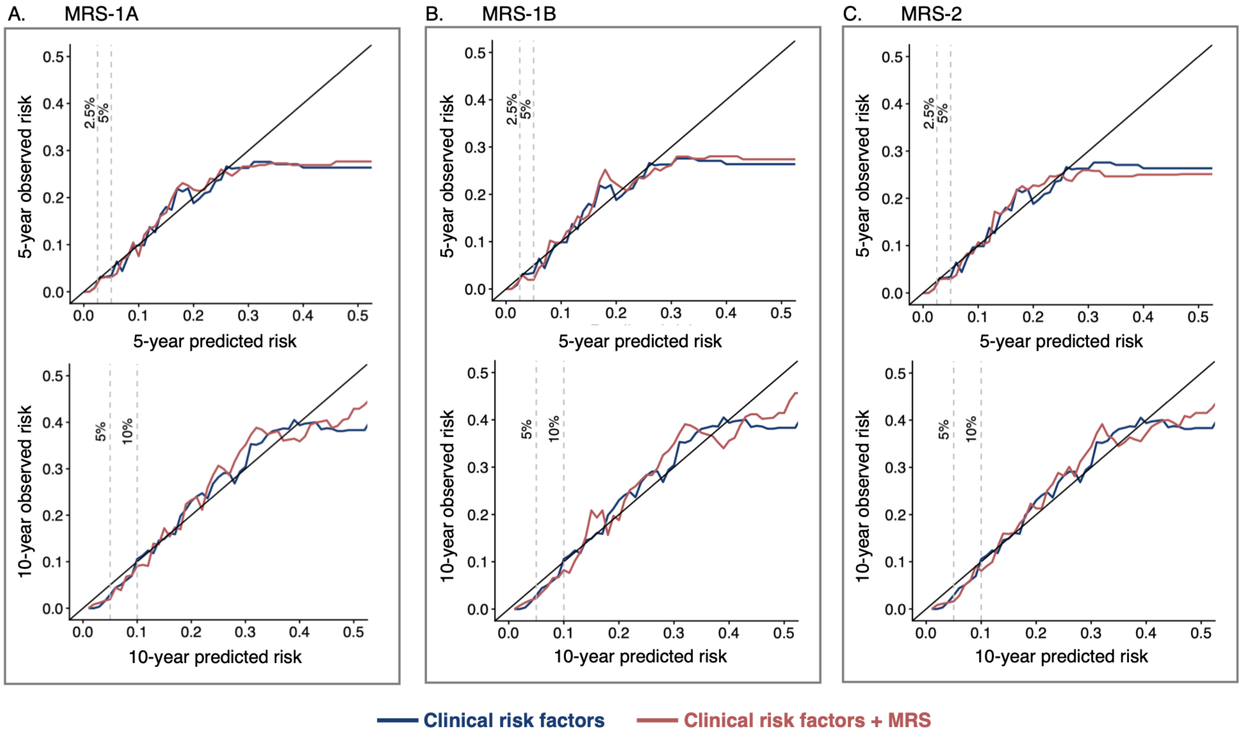


**Figure S10. Calibration of ASCVD risk prediction models with and without methylation risk scores (MRS).** Calibration plots comparing predicted versus observed risk for atherosclerotic cardiovascular disease (ASCVD) at 5 years (top row) and 10 years (bottom row). Panels A-C correspond to models incorporating different methylation risk scores (MRS-1A, MRS-1B, and MRS-2, respectively). Blue lines represent models including clinical risk factors only, while red lines represent models including clinical risk factors + MRS. The diagonal line indicates perfect calibration. Dashed vertical lines mark clinically relevant percentiles of predicted risk. Clinical risk factors include body mass index, smoking status, HDL-C, LDL-C, systolic blood pressure, lipid-lowering medication use, antihypertensive medication use, and diabetes history. All models were adjusted for age, sex, genetic principal components 1-10, and estimated blood cell proportions.

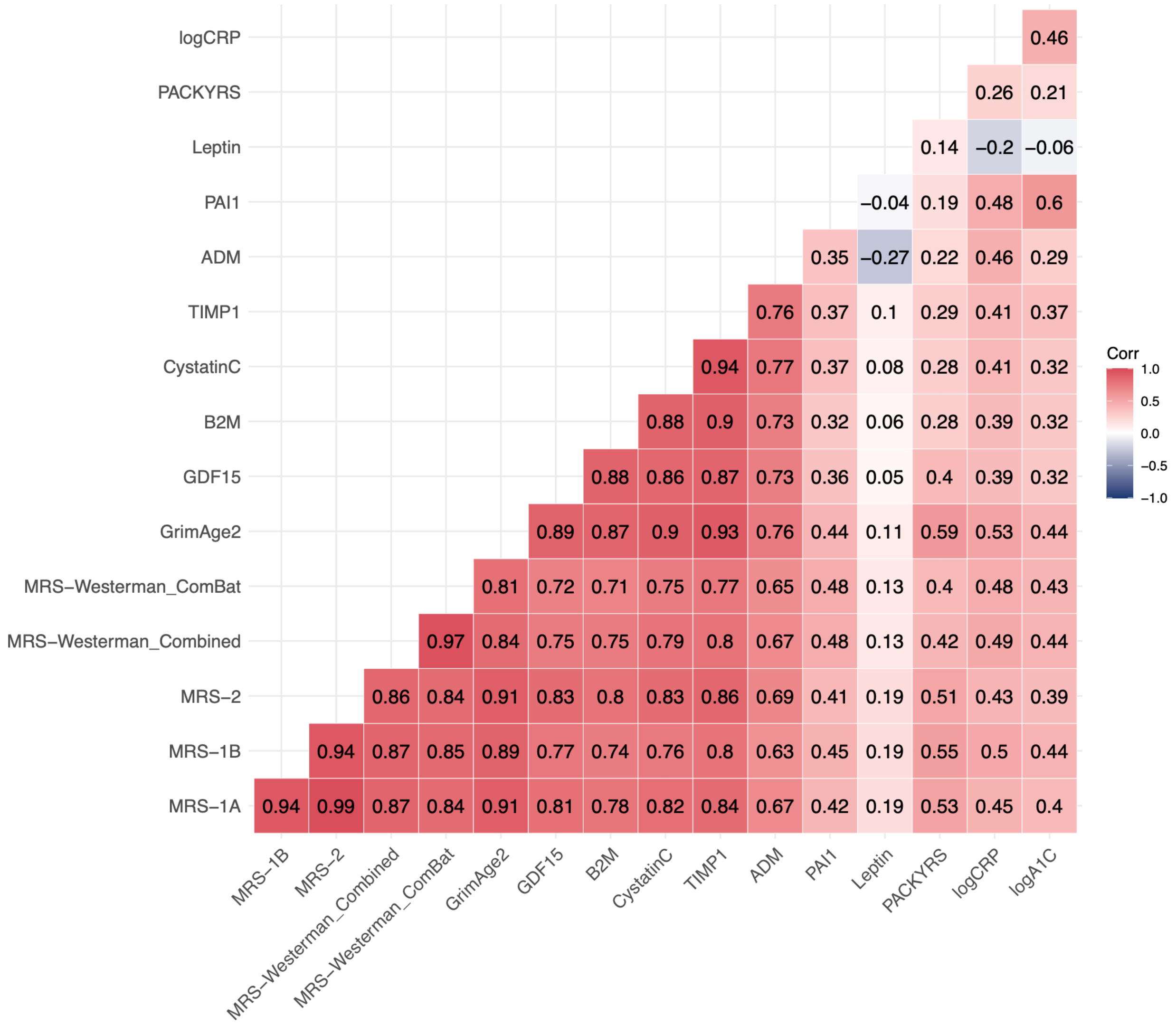


**Figure S11. Pairwise correlations among GrimAge2, GrimAge2 subcomponents, and ASCVD- and CAD-specific methylation risk scores (MRS).** Pairwise Pearson correlation coefficients are shown for the three MVP-derived ASCVD MRS (MRS-1A, MRS-1B, and MRS-2), the two previously published CAD-specific MRS developed by Westerman et al. (Combined and ComBat), GrimAge2, and the 10 GrimAge2 DNA methylation-based surrogate components (growth differentiation factor 15 [GDF15], β2-microglobulin [B2M], cystatin C, tissue inhibitor of metalloproteinases 1 [TIMP1], adrenomedullin [ADM], plasminogen activator inhibitor 1 [PAI1], leptin, smoking pack-years [PACKYRS], C-reactive protein [logCRP], and hemoglobin A1c [logA1C]).

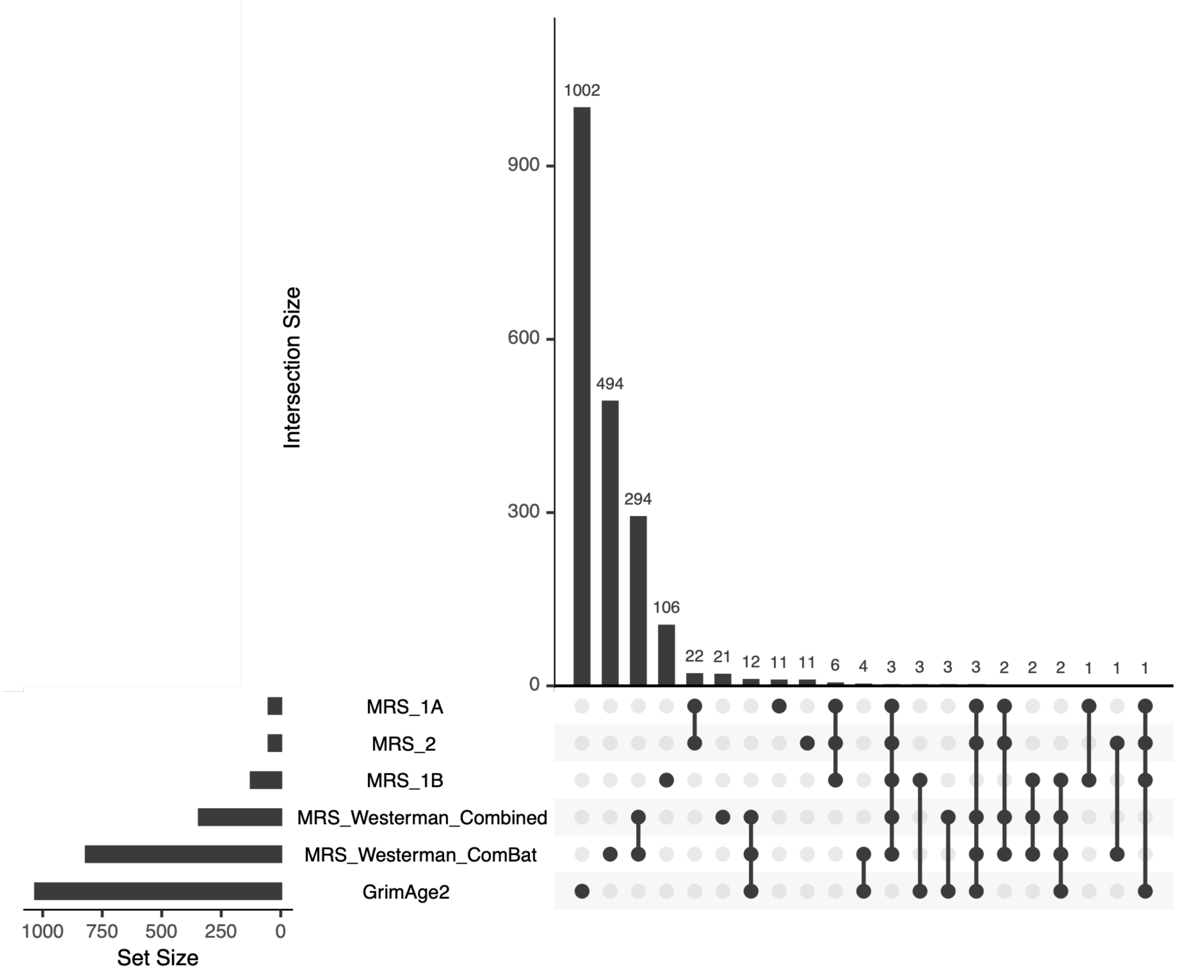


**Figure S12. Overlap in CpGs among GrimAge2 and ASCVD- and CAD-specific methylation risk scores (MRS).** UpSet plot showing the overlap of CpGs among the three MVP-derived ASCVD MRS (MRS-1A, MRS-1B, and MRS-2), the two previously published CAD-specific MRS developed by Westerman et al. (Combined and ComBat), and GrimAge2. Horizontal bars indicate the total number of CpGs in each score (set size), and vertical bars indicate the number of CpGs shared across the combinations of scores denoted by the connected dots below.

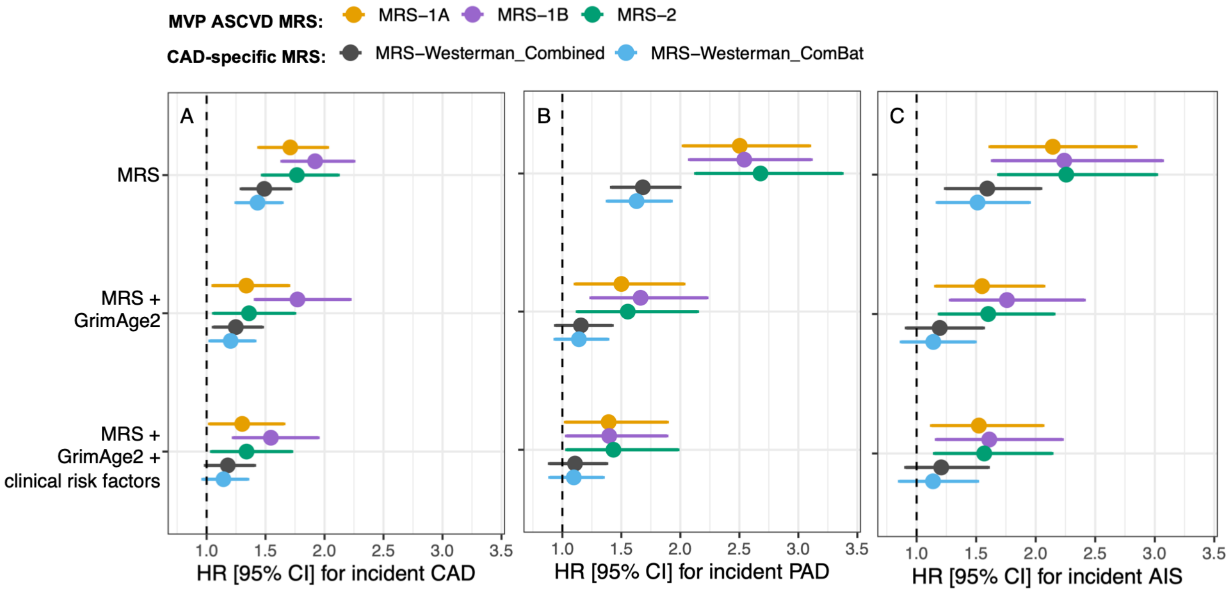


**Figure S13. Associations of methylation risk scores (MRS) with incident CAD, PAD, or AIS with and without adjustment for GrimAge2 and clinical risk factors.** Hazard ratios (HRs) and 95% confidence intervals from Cox proportional hazards models for coronary artery disease (CAD) (A), peripheral artery disease (PAD) (B), and acute ischemic stroke (AIS) (C). The number of cases was 489 for CAD, 285 for PAD, and 67 for AIS in this study. HR [95%CI] are shown per 1-SD increase in each MRS. Clinical risk factors included body mass index, smoking status, HDL-C, LDL-C, systolic blood pressure, lipid-lowering medication use, antihypertensive medication use, and diabetes history. All models were adjusted for age, sex, genetic principal components 1-10, and estimated blood cell proportions. Due to the smaller number of AIS cases, models for AIS were adjusted for a reduced set of covariates (age, genetic principal components 1-5, and estimated blood cell proportions).

**Supplemental Text S1. VA Million Veteran Program core acknowledgements list.**

**VA Million Veteran Program**
**Core Acknowledgements for Publications**
**June 2026**

**MVP Program Office**

- Sumitra Muralidhar, Ph.D., Program Director
  US Department of Veterans Affairs, 810 Vermont Avenue NW, Washington, DC 20420
- Jennifer Moser, Ph.D., Associate Director, Scientific Programs
  US Department of Veterans Affairs, 810 Vermont Avenue NW, Washington, DC 20420
- Jennifer E. Deen, B.S., Associate Director, Cohort & Public Relations
  US Department of Veterans Affairs, 810 Vermont Avenue NW, Washington, DC 20420

**MVP Steering Committee**

- Co-Chair: J. Michael Gaziano, M.D., M.P.H.
  VA Boston Healthcare System, 150 S. Huntington Avenue, Boston, MA 021
- Co-Chair: Dave Oslin, M.D.
  Philadelphia VA Medical Center, 3900 Woodland Avenue, Philadelphia, PA 19104
- Sumitra Muralidhar, Ph.D., Ex-Officio
  US Department of Veterans Affairs, 810 Vermont Avenue NW, Washington, DC 20420
- Drew Helmer, M.D., M.S.
  Michael E. DeBakey VA Medical Center, 2002 Holcombe Boulevard, Houston, TX 77030
- Adriana Hung, M.D., M.P.H.
  VA Tennessee Valley Healthcare System, 1310 24th Avenue, South Nashville, TN 37212
- Philip S. Tsao, Ph.D.
  VA Palo Alto Health Care System, 3801 Miranda Avenue, Palo Alto, CA 94304
- Deepak Voora, M.D.
  Durham VA Medical Center, 508 Fulton Street, Durham, NC 27705

**MVP Co-Principal Investigators**

- J. Michael Gaziano, M.D., M.P.H.
  VA Boston Healthcare System, 150 S. Huntington Avenue, Boston, MA 02130
- Philip S. Tsao, Ph.D.
  VA Palo Alto Health Care System, 3801 Miranda Avenue, Palo Alto, CA 94304

**MVP Core Operations**

- Jessica V. Brewer, M.P.H., Director, MVP Cohort Operations
  VA Boston Healthcare System, 150 S. Huntington Avenue, Boston, MA 02130
- Kelly Cho, M.P.H, Ph.D., Director, MVP Phenomics
  VA Boston Healthcare System, 150 S. Huntington Avenue, Boston, MA 02130
- Lori Churby, B.S., Director, MVP Regulatory Affairs
  VA Palo Alto Health Care System, 3801 Miranda Avenue, Palo Alto, CA 94304

- Yonghui Jia, Ph.D., Director, VA Central Biorepository
  VA Boston Healthcare System, 150 S. Huntington Avenue, Boston, MA 02130
- Jacob T. Kean, Ph.D., Acting Director, VA Informatics and Computing Infrastructure (VINCI)
  VA Salt Lake City Health Care System, 500 Foothill Drive, Salt Lake City, UT 84148
- Saiju Pyarajan Ph.D., Director, Data and Computational Sciences
  VA Boston Healthcare System, 150 S. Huntington Avenue, Boston, MA 02130
- Robert Ringer, Pharm.D., Director, VA Albuquerque Central Biorepository
  New Mexico VA Health Care System, 1501 San Pedro Drive SE, Albuquerque, NM 87108
- Luis E. Selva, Ph.D., Director, MVP Biorepository Coordination
  VA Boston Healthcare System, 150 S. Huntington Avenue, Boston, MA 02130
- Shahpoor (Alex) Shayan, M.S., Director, MVP PRE Informatics
  VA Boston Healthcare System, 150 S. Huntington Avenue, Boston, MA 02130
- Brady Stephens, M.S., Principal Investigator, MVP Information Center
  Canandaigua VA Medical Center, 400 Fort Hill Avenue, Canandaigua, NY 14424
- Stacey B. Whitbourne, Ph.D., Director, MVP Cohort Development and Management
  VA Boston Healthcare System, 150 S. Huntington Avenue, Boston, MA 02130
